# Associations between residential exposure to volatile organic compounds (VOCs) and liver injury markers

**DOI:** 10.1101/2021.08.25.21262640

**Authors:** Banrida Wahlang, Tyler C. Gripshover, Hong Gao, Tatiana Krivokhizhina, Rachel J. Keith, Israel D. Sithu, Shesh N. Rai, Aruni Bhatnagar, Craig J. McClain, Sanjay Srivastava, Mathew C. Cave

## Abstract

Occupational exposures to volatile organic compounds (VOCs) have been associated with numerous health complications including steatohepatitis and liver cancer. However, the potential impact of environmental/residential VOC exposures on liver health and function is largely unknown. To address this knowledge gap, the objective of this cross-sectional study is to investigate associations between VOCs and liver injury biomarkers in community residents. Subjects were recruited from six Louisville neighborhoods, and informed consent was obtained. Exposure biomarkers included 16 creatinine-adjusted urinary metabolites corresponding to 12 parent VOCs. Serological disease biomarkers measured included cytokertain-18 (K18 M65 and M30), liver enzymes and direct bilirubin. Associations between exposure and disease biomarkers were assessed using generalized linear models. Smoking status was confirmed through urinary cotinine levels. The population comprised of approximately 60% females and 40% males; White persons accounted 78% of the population; with more nonsmokers (n=413) than smokers (n=250). When compared to nonsmokers, Males (45%) and Black persons (26%) were more likely to be smokers. In the overall population, metabolites of acrolein, acrylonitrile, acrylamide, 1,3-butadiene, crotonaldehyde, styrene and xylene were positively associated with alkaline phosphatase (ALP). These associations persisted in smokers, with the exception of crotonaldehyde, and addition of N,N dimethylformamide and propylene oxide metabolites. Although no positive associations were observed for K18 M30, the benzene metabolite was positively associated with bilirubin, irrespective of smoking status. Taken together, the results demonstrated that selected VOCs were positively associated with liver injury biomarkers. These findings will enable better risk assessment and identification of populations vulnerable to liver disease.

## INTRODUCTION

Volatile organic compounds (VOCs) are environmental chemicals that are anthropogenic or naturally-occurring; these chemicals have a high vapor pressure and low water solubility, and therefore, are often emitted as gases in indoor and outdoor air. While most of these volatile chemicals, such as trichlorethylene, toluene and benzene, are typically used as solvents in a wide range of industrial and chemical applications; other VOCs including acrylamide and styrene have been used in rubber and plastic manufacturing (Cruz and Bowen, 2021). Their applications in a variety of industrial products, such as petroleum fuels and hydraulic liquids, also make them common ground-water contaminants. Importantly, VOCs are also emitted from numerous business and household products (Lim *et al*., 2014), as well as from cigarette/tobacco smoke (Nazaroff and Singer, 2004; Sheu *et al*., 2020), resulting in different kinds of exposures extending from occupational or industrial to residential or environmental.

Occupational exposures to VOCs have been shown to cause adverse health effects, ranging from different forms of cancers such as lung, liver, and kidney cancers, to neurotoxicity and neuro-behavioral deficits, depending on the type(s) of VOC (Cave *et al*., 2010; Cruz and Bowen, 2021; Lamplugh *et al*., 2019; Walker *et al*., 2016; Warden *et al*., 2018). These occupational exposures oftentimes occur at higher doses and can be acute (short-term exposures) or chronic (longer-term exposures). In contrast, residential VOC exposures often occur at lower doses over an extended period of time, usually when people inhale VOCs residing within their environment, which frequently are VOCs found in household or indoor air (Vardoulakis *et al*., 2020). Although occupational VOC exposures have been associated with cardiovascular diseases and liver injury (Cave *et al*., 2011; Cave, et al., 2010; Gresner *et al*., 2021; Ma *et al*., 2010), less is known about the impact of residential VOC exposures on cardio-metabolic diseases including liver injury and non-alcoholic fatty liver disease (NAFLD).

The liver is the principal organ for chemical detoxication, which also makes it a general target organ for many environmental chemicals. Indeed, previous epidemiologic and rodent studies have demonstrated the toxicological effects of VOCs on liver health and function (Lang and Beier, 2018; Wahlang *et al*., 2019). Clinically, liver injury has traditionally been assessed by circulating liver enzyme levels such as plasma alanine and aspartate transaminases (ALT, AST), and alkaline phosphatase (ALP). More recently, our group and others have used plasma cytokeratin 18 (K18) M65 and M30 as alternative non-invasive biomarkers for liver injury, in relation to environmental chemical exposures (Cave, et al., 2011; Clair *et al*., 2018; Werder *et al*., 2020). Aside from elevated K18 levels, exposures to industrial VOCs, such as vinyl chloride and styrene, have been associated with toxicant-associated steatohepatitis (TASH) in rubber plant workers, with mechanistic toxicity including necrosis (Cave, et al., 2011; Cave, et al., 2010). TASH is a form of non-alcoholic steatohepatitis (NASH), and share similar pathologies and molecular disease development events as NASH (Wahlang *et al*., 2013). Both TASH and NASH fall under the bigger description of liver pathology, namely, NAFLD, which consist of a spectrum of hepatic disorders, initially manifested by fat accumulation in the liver (steatosis), and this could be accompanied by inflammation (steatohepatitis). If untreated, steatohepatitis could progress to fibrosis and cirrhosis and fulminant liver failure. Notably, the prevalence of TASH has also been observed in populations exposed to other classes of pollutants such as persistent organic pollutants (Clair, et al., 2018; Wahlang, et al., 2013; Wahlang, et al., 2019), and therefore it is important to assess the effects of residential VOCs on such liver endpoints.

Health consequences resulting from exposures to environmental pollutants are heavily dictated by non-pollutant factors such as proximity to a polluted site, socio-economic factors, race, sex and lifestyle choices including smoking, high caloric intake and physical activity. These factors influence levels of both exposure and disease biomarkers, as well as health outcomes associated with exposures; for instance smokers may have higher VOC exposure levels originating from cigarette smoke (Nazaroff and Singer, 2004). Most environmental epidemiologic studies have begun taken into consideration the role of such factors when investigating associations and correlations between environmental chemical exposure and disease. In concordance with such studies, the objective of the current study is to investigate associations between residential exposures to VOCs using urinary VOC metabolites, and liver disease biomarkers, to better understand hepatic mechanisms related to chronic, low dose VOC inhalation. In addition, the study utilizes a stratified approach to better assess differences in associations between smokers *vs.* non-smokers, as well as how sex and race may influence the observed associations.

## MATERIALS AND METHOD

### Recruitment and Study Population

The current study which is part of the <u>H</u>ealth, <u>E</u>nvironment and <u>A</u>ction in <u>L</u>ouisville (HEAL) study, was approved by the University of Louisville Institutional Review Board. A total of 735 consenting study subjects were enrolled by the University of Louisville, from six neighborhoods in South Louisville, Kentucky, between May 2018 and September 2019. All subjects were between 25 to 70 years of age. Individuals unwilling or unable to provide written informed consent were excluded. Individuals with significant and/or severe comorbidities were also excluded. These comorbidities included HIV/AIDS, active treatment for cancer, and active bleeding including wounds. Other excluded subjects were pregnant women, prisoners, or other vulnerable populations.

Blood and urine specimens were collected from the enrolled subjects. Demographic information including age, sex, race, smoking and drinking history, medical information including diabetes, cardiovascular diseases, and other chronic conditions were obtained through the questionnaires. BMI was calculated from the self-reported height and weight. Demographic characteristics of the study population are provided in **Table 1**.

**Table 1.**
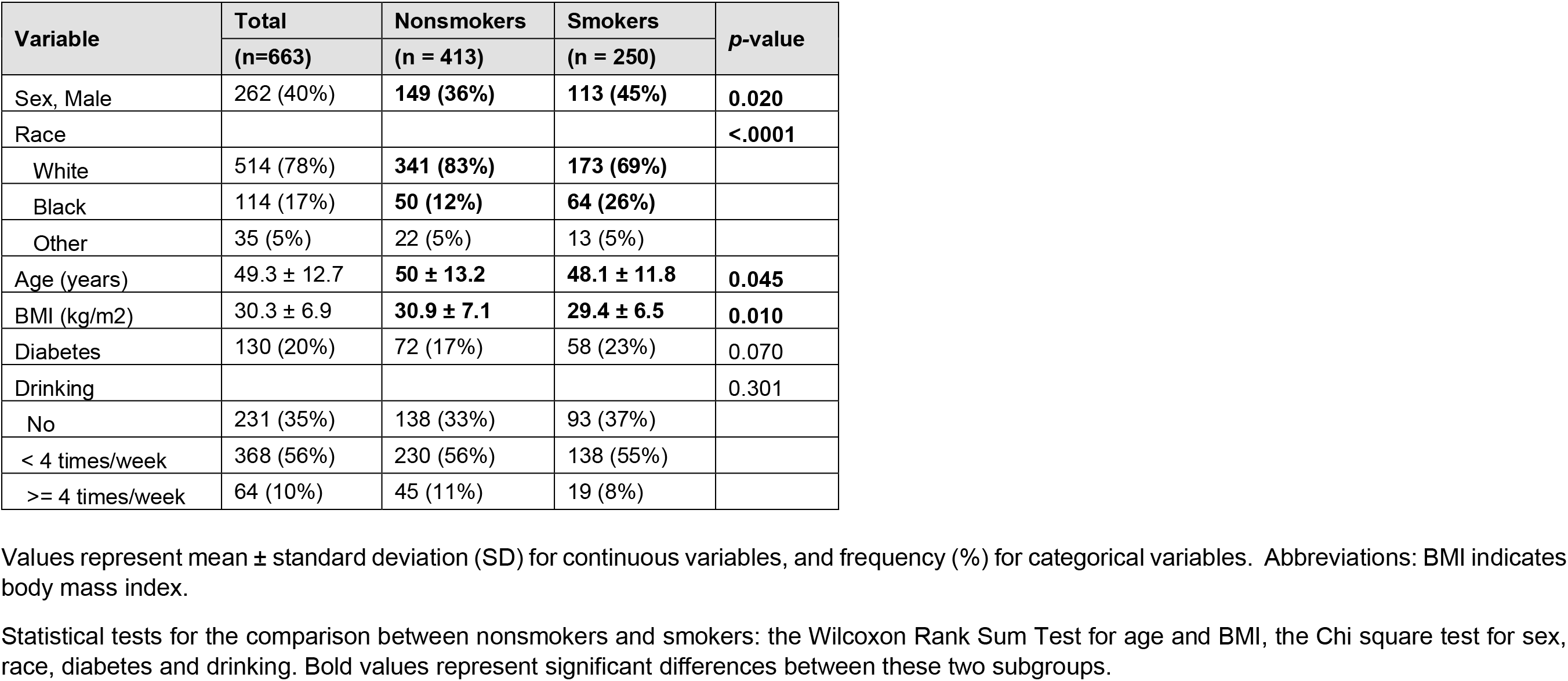
Characteristics of the study population by nonsmokers and smokers.

Of those enrolled in the current study, 46 subjects did not provide blood specimens, 2 subjects did not have liver enzymes/bilirubin measurements, 10 subjects did not provide urine specimens, 11 subjects were missing covariates such as BMI or drinking information, and 2 subjects had severe obesity. One subject with an extreme value of serum cytokeratin 18 (K18) M30/M65 ratio was also excluded from the analysis. Smoking status was confirmed using normalized urinary cotinine levels, with a value of >40 µg/g creatinine indicating current smoking status. The final study population consisted of 663 subjects (**Figure 1**)

**Figure 1.**
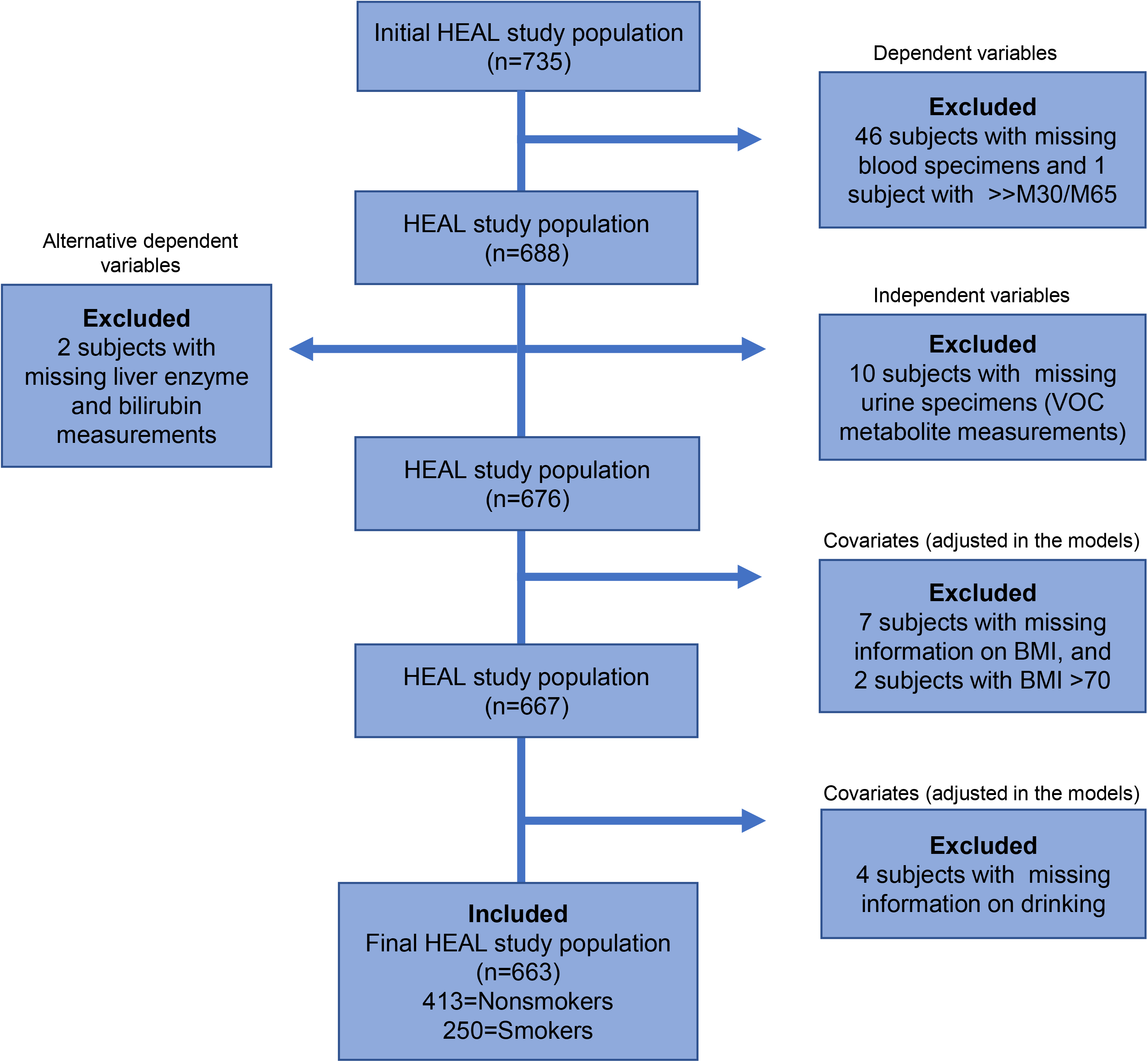
A flowchart depicting the final study population after applying exclusion criteria.

### Quantification of Urinary VOC Metabolites

A panel of 16 VOC metabolites from 12 parent compounds, and the nicotine metabolite (cotinine) in spot urine samples, were quantified using a modified version of ultra-performance liquid chromatography-mass spectrometry (UPLC-MS/MS) method (Alwis *et al*., 2012; Lorkiewicz *et al*., 2019). All analytes were normalized to urinary creatinine levels to adjust for dilution. A list of all 16 VOC metabolites and their abbreviations, the corresponding parent compounds, and the percent lower than the limit of detection (% < LOD) is provided in **Table 2**.

**Table 2.**
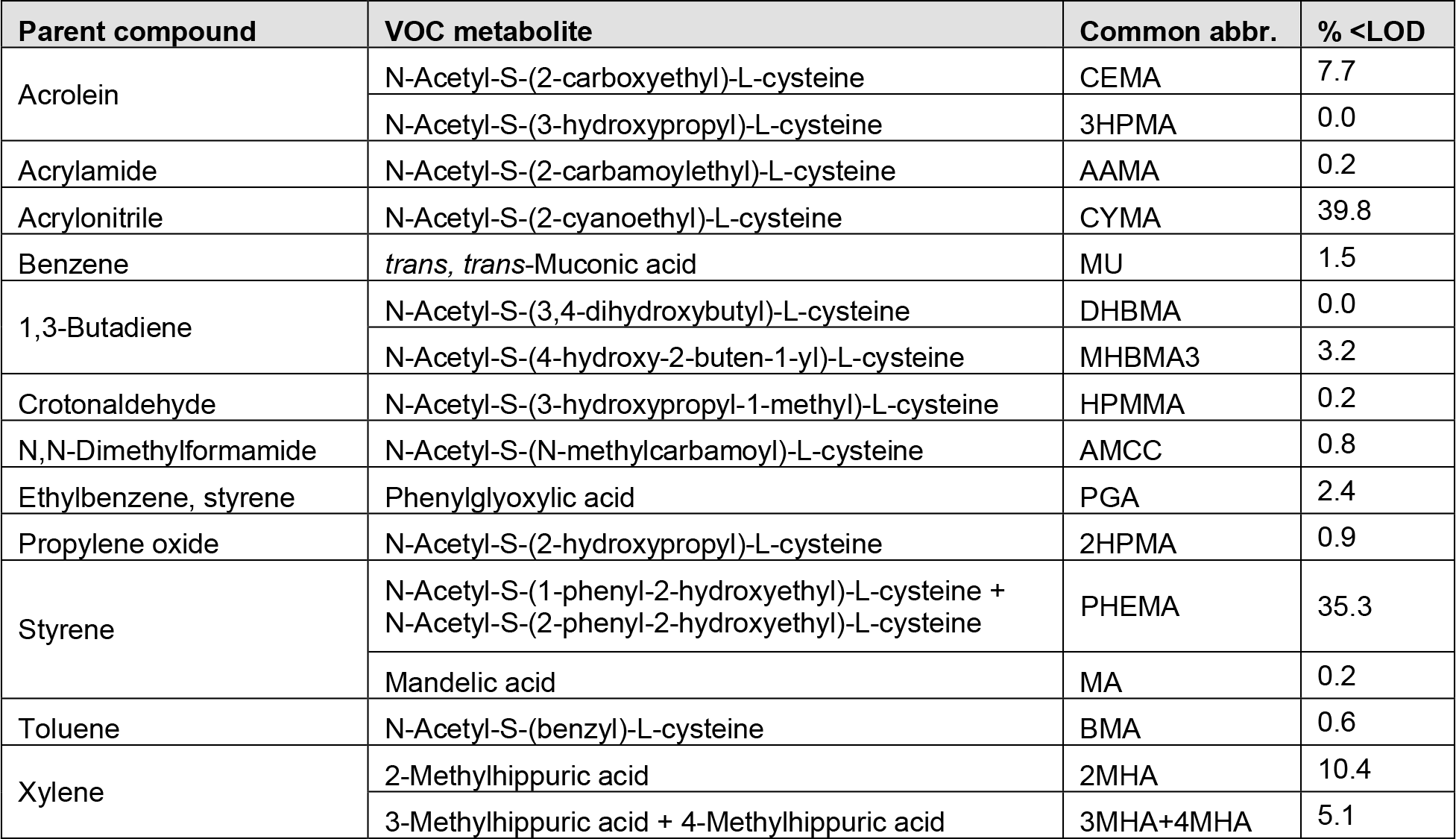
A list of the 16 volatile organic compounds (VOC) metabolites measured in the total study cohort, their parent compounds, and the percent lower than the limit of detection (% <LOD).

| Parent compound | VOC metabolite | Common abbr. | % <LOD |
| --- | --- | --- | --- |
| Acrolein | N-Acetyl-S-(2-carboxyethyl)-L-cysteine | CEMA | 7.7 |
|  | N-Acetyl-S-(3-hydroxypropyl)-L-cysteine | 3HPMA | 0.0 |
| Acrylamide | N-Acetyl-S-(2-carbamoyl-ethyl)-L-cysteine | AAMA | 0.2 |
| Acrylonitrile | N-Acetyl-S-(2-cyanoethyl)-L-cysteine | CYMA | 39.8 |
| Benzene | *trans, trans*-Muconic acid | MU | 1.5 |
| 1,3-Butadiene | N-Acetyl-S-(3,4-dihydroxybutyl)-L-cysteine | DHBMA | 0.0 |
|  | N-Acetyl-S-(4-hydroxy-2-buten-1-yl)-L-cysteine | MHBMA3 | 3.2 |
| Crotonaldehyde | N-Acetyl-S-(3-hydroxypropyl-1-methyl)-L-cysteine | HPMMA | 0.2 |
| N,N-Dimethylformamide | N-Acetyl-S-(N-methylcarbamoyl)-L-cysteine | AMCC | 0.8 |
| Ethylbenzene, styrene | Phenylglyoxylic acid | PGA | 2.4 |
| Propylene oxide | N-Acetyl-S-(2-hydroxypropyl)-L-cysteine | 2HPMA | 0.9 |
| Styrene | N-Acetyl-S-(1-phenyl-2-hydroxyethyl)-L-cysteine +<br>N-Acetyl-S-(2-phenyl-2-hydroxyethyl)-L-cysteine | PHEMA | 35.3 |
|  | Mandelic acid | MA | 0.2 |
| Toluene | N-Acetyl-S-(benzyl)-L-cysteine | BMA | 0.6 |
| Xylene | 2-Methylhippuric acid | 2MHA | 10.4 |
|  | 3-Methylhippuric acid + 4-Methylhippuric acid | 3MHA+4MHA | 5.1 |

### Quantification of Serum Cytokeratin 18 (K18), Liver Enzymes and Bilirubin

Serum K18 M65 and M30 levels (U/L) were measured using the PEVIVA M65 (Product No. 10011) and PEVIVA M30 Apoptosense (Product No. 10020) ELISA kits (Diapharma Group Inc., West Chester, OH), according to the manufacturers’ instructions. K18 is an intermediate filament protein highly expressed by the liver, and activation of cell death pathways lead to cleavage of K18 into caspase-cleaved M30 fragments, indicative of apoptosis, as opposed to intact K18 M65, indicative of necrosis. Serum liver enzymes, namely ALT, AST, and ALP and direct bilirubin were measured on Ace Axcel® Clinical Chemistry System (Alfa Wassermann, West Caldwell, NJ), according to the manufacturers’ instructions.

### Statistical Analyses

The characteristics of the total study population (n = 663), nonsmokers (n = 413), and smokers (n = 250), are expressed as mean ± standard deviation (SD) for continuous variables, and frequency (%) for categorical variables. To examine the different characteristics between nonsmokers and smokers, the Wilcoxon Rank Sum Test was conducted for age and BMI, and the Chi square test was conducted for sex, race, diabetes and drinking (**Table 1**).

Descriptive statistics were conducted for all the independent variables (16 VOC metabolites) and dependent variables (7 liver biomarkers). Since the distributions of all these variables were right skewed, they were log transformed to improve the normality when *t*-tests were performed between nonsmokers and smokers. Pearson correlations were performed across 16 log-transformed urinary VOC metabolites and 7 log-transformed liver biomarkers in the total study population (n = 663); nonsmokers (n=413) and smokers (n=250) (**Figure 2A-C** of correlation heat maps). To examine the associations between VOC exposures and liver biomarkers, we constructed generalized linear models by regressing each of the 16 VOC metabolites against each of the 7 liver biomarkers in the total study population (**Table 4**). Both independent variables (VOC metabolites) and dependent variables (liver biomarkers) were log transformed to improve the normality in the models. Models were adjusted for 6 common liver-related confounders: age, sex, race, BMI, diabetes status and alcohol consumption. During the model selection process, we found that the effect of VOC metabolites on ALP might be mediated by smoking. Therefore, we stratified the analyses by smoking status for all the models. In addition, we compared the adjusted associations between urinary levels of 16 VOC metabolites and two liver biomarkers (ALP and Bilirubin) in the total study population (n=663) between 6 adjustments and 7 adjustments (6 adjustments + smoking) as shown in **Supplemental Table S3 and S4 and Figure S1 and S2**. The statistical significance was set at the *p*-value <0.05. All statistical analyses were performed using SAS, version 9.4 (SAS Institute, Inc., Cary, North Carolina). The heat maps were produced in R (version 4.0.5) and the forest plots were produced in Graph Pad Prism, version 9.1 (Graph Pad Software, La Jolla, California).

**Figure 2.**
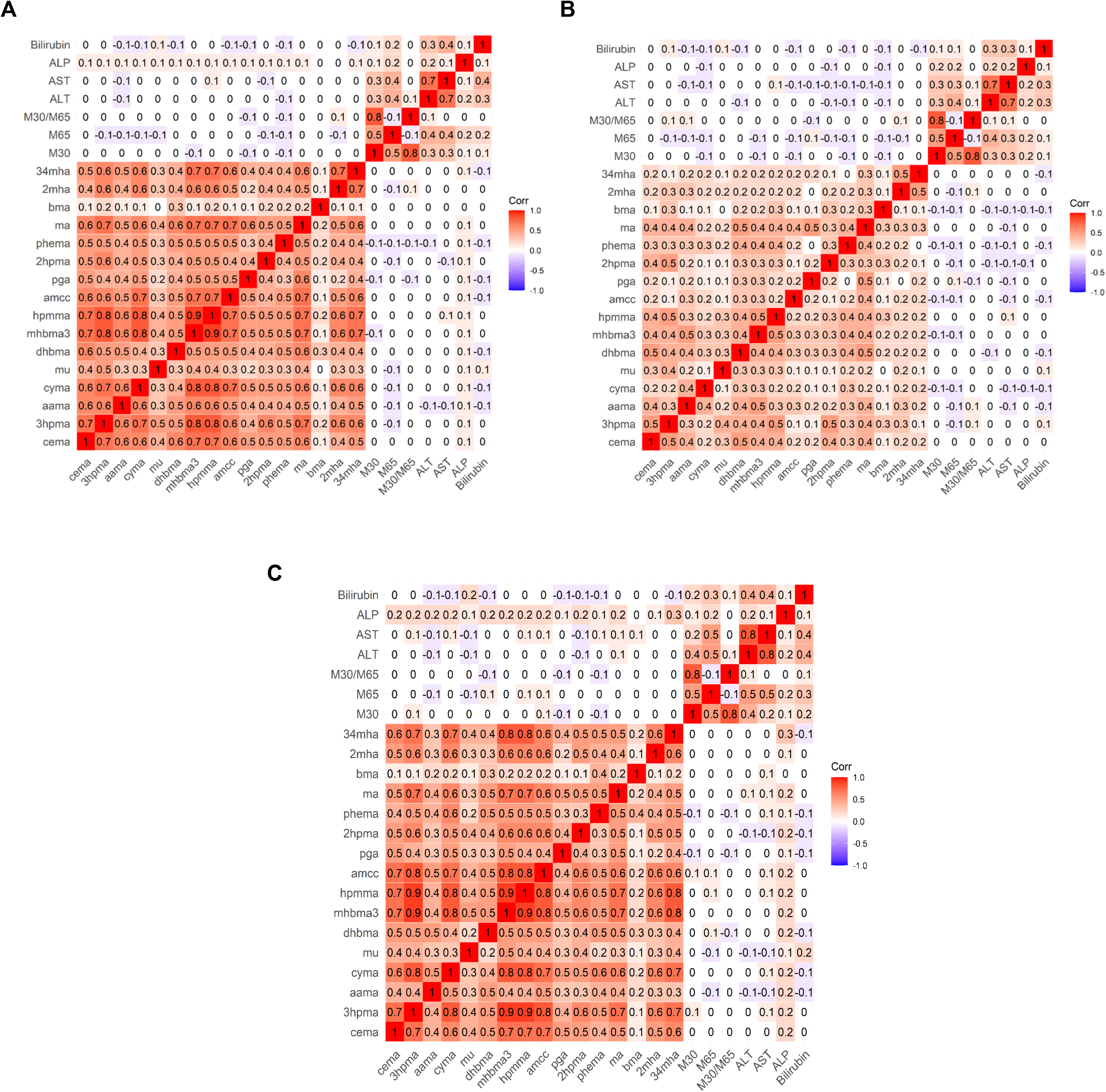
A heatmap of Pearson correlation coefficients between 16 log-transformed urinary VOC metabolites and 7 log-transformed liver biomarkers in A) the total study population (n = 663), B) nonsmokers (n = 413), and C) smokers (n=250).

**Table 3.**
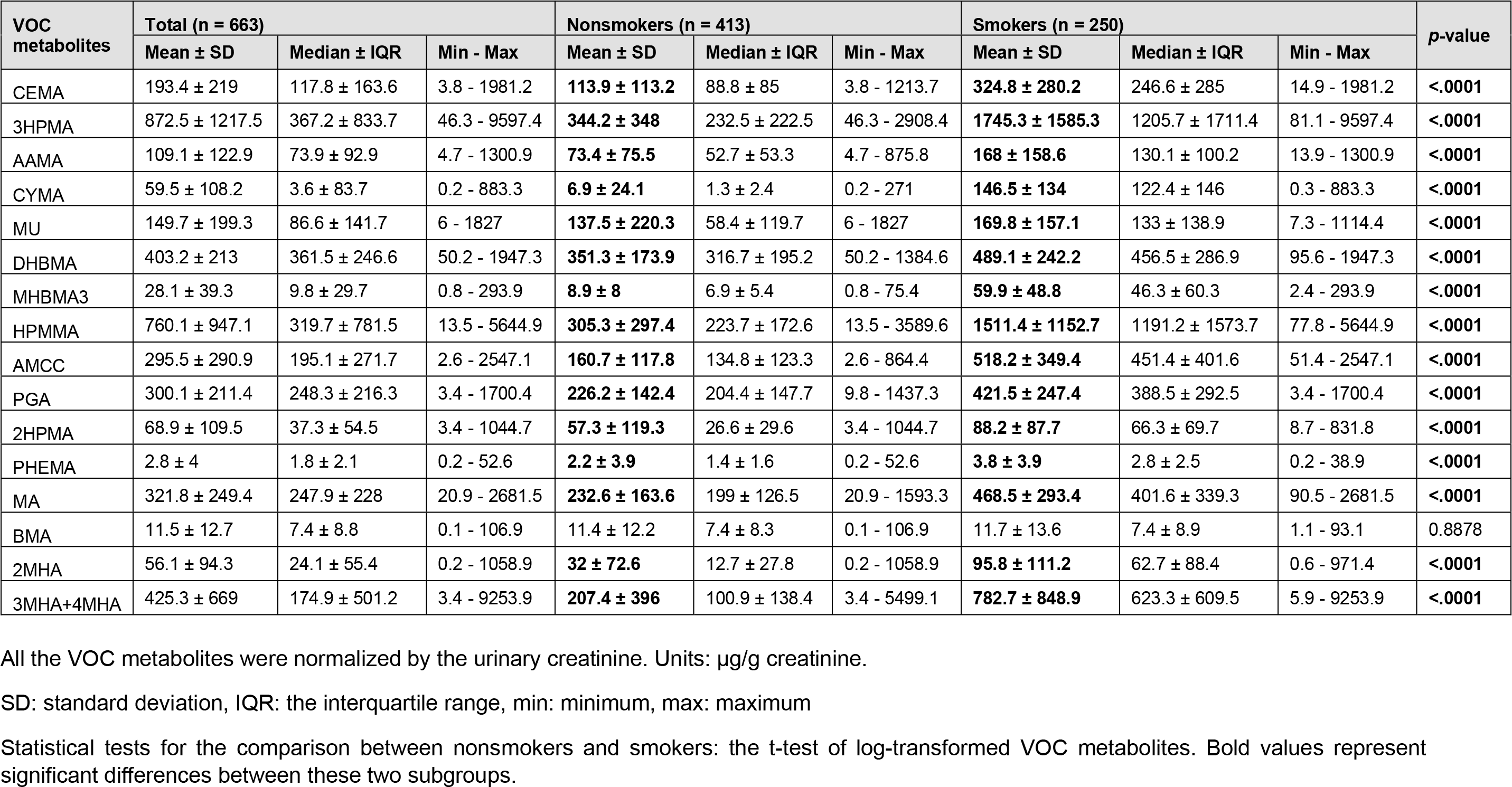
Distribution of VOC metabolites within nonsmokers and smokers.

**Table 4.** Relationships between the 16 VOC metabolites and 7 liver biomarkers in the total study population (n=663).

| VOC metabolites | Keratin 18 (n = 663) |  |  |  |  |  | Enzymes and Bilirubin (n = 663) |  |  |  |  |  |  |  |
| --- | --- | --- | --- | --- | --- | --- | --- | --- | --- | --- | --- | --- | --- | --- |
|  | M30 |  | M65 |  | M30/M65 |  | ALT |  | AST |  | ALP |  | Bilirubin |  |
| | $\beta \pm SE$ | p-value | $\beta \pm SE$ | p-value | $\beta \pm SE$ | p-value | $\beta \pm SE$ | p-value | $\beta \pm SE$ | p-value | $\beta \pm SE$ | p-value | $\beta \pm SE$ | p-value |
| CEMA | -0.01 ± 0.04 | 0.729 | -0.02 ± 0.02 | 0.475 | 0 ± 0.04 | 0.938 | 0.04 ± 0.03 | 0.177 | 0 ± 0.02 | 0.944 | **0.03 ± 0.01** | **0.004** | -0.01 ± 0.02 | 0.532 |
| 3HPMA | 0.02 ± 0.04 | 0.652 | -0.02 ± 0.02 | 0.267 | 0.04 ± 0.03 | 0.219 | 0.01 ± 0.02 | 0.627 | 0.01 ± 0.02 | 0.425 | **0.03 ± 0.01** | **0.011** | 0.01 ± 0.02 | 0.451 |
| AAMA | 0.02 ± 0.05 | 0.684 | -0.03 ± 0.03 | 0.311 | 0.05 ± 0.04 | 0.264 | -0.02 ± 0.03 | 0.421 | -0.04 ± 0.02 | 0.062 | **0.05 ± 0.01** | **0** | **-0.06 ± 0.02** | **0.008** |
| CYMA | 0 ± 0.02 | 0.811 | 0 ± 0.01 | 0.703 | 0.01 ± 0.02 | 0.606 | 0 ± 0.01 | 0.685 | 0 ± 0.01 | 0.946 | **0.01 ± 0** | **0.012** | -0.01 ± 0.01 | 0.08 |
| MU | -0.05 ± 0.04 | 0.188 | **-0.07 ± 0.02** | **0.004** | 0.01 ± 0.04 | 0.695 | -0.01 ± 0.02 | 0.572 | -0.01 ± 0.02 | 0.466 | 0.01 ± 0.01 | 0.582 | **0.05 ± 0.02** | **0.003** |
| DHBMA | -0.04 ± 0.08 | 0.638 | 0.03 ± 0.05 | 0.582 | -0.06 ± 0.07 | 0.376 | -0.04 ± 0.05 | 0.463 | -0.02 ± 0.03 | 0.481 | 0.04 ± 0.02 | 0.083 | **-0.09 ± 0.04** | **0.014** |
| MHBMA3 | -0.02 ± 0.04 | 0.559 | -0.01 ± 0.02 | 0.803 | -0.02 ± 0.03 | 0.624 | 0.01 ± 0.02 | 0.506 | 0.01 ± 0.01 | 0.556 | **0.03 ± 0.01** | **0.004** | -0.01 ± 0.02 | 0.676 |
| HPMMA | 0.02 ± 0.04 | 0.538 | 0.01 ± 0.02 | 0.819 | 0.02 ± 0.04 | 0.59 | 0.04 ± 0.02 | 0.117 | 0.03 ± 0.02 | 0.051 | **0.02 ± 0.01** | **0.028** | -0.01 ± 0.02 | 0.585 |
| AMCC | -0.04 ± 0.04 | 0.352 | -0.03 ± 0.03 | 0.279 | -0.01 ± 0.04 | 0.739 | 0.03 ± 0.03 | 0.325 | -0.01 ± 0.02 | 0.441 | 0.02 ± 0.01 | 0.15 | **-0.05 ± 0.02** | **0.007** |
| PGA | -0.08 ± 0.05 | 0.138 | 0.04 ± 0.03 | 0.194 | **-0.12 ± 0.05** | **0.012** | 0.02 ± 0.03 | 0.564 | -0.02 ± 0.02 | 0.375 | 0.02 ± 0.01 | 0.091 | -0.04 ± 0.02 | 0.07 |
| 2HPMA | -0.02 ± 0.04 | 0.681 | -0.03 ± 0.03 | 0.194 | 0.01 ± 0.04 | 0.698 | -0.02 ± 0.03 | 0.488 | -0.02 ± 0.02 | 0.164 | 0.02 ± 0.01 | 0.087 | -0.01 ± 0.02 | 0.586 |
| PHEMA | -0.08 ± 0.05 | 0.088 | -0.05 ± 0.03 | 0.089 | -0.03 ± 0.04 | 0.429 | -0.03 ± 0.03 | 0.339 | 0 ± 0.02 | 0.867 | 0.02 ± 0.01 | 0.104 | -0.04 ± 0.02 | 0.067 |
| MA | -0.01 ± 0.06 | 0.886 | 0 ± 0.04 | 0.979 | -0.01 ± 0.06 | 0.886 | 0.05 ± 0.04 | 0.196 | 0 ± 0.03 | 0.977 | **0.05 ± 0.02** | **0.005** | -0.03 ± 0.03 | 0.351 |
| BMA | -0.01 ± 0.05 | 0.785 | -0.02 ± 0.03 | 0.612 | 0 ± 0.05 | 0.979 | 0.01 ± 0.03 | 0.853 | 0 ± 0.02 | 0.947 | -0.02 ± 0.01 | 0.255 | 0 ± 0.02 | 0.869 |
| 2MHA | 0 ± 0.03 | 0.996 | **-0.04 ± 0.01** | **0.011** | 0.04 ± 0.02 | 0.095 | 0 ± 0.02 | 0.77 | 0 ± 0.01 | 0.659 | 0.01 ± 0.01 | 0.293 | -0.01 ± 0.01 | 0.376 |
| 3MHA+4MHA | -0.01 ± 0.03 | 0.774 | 0 ± 0.02 | 0.859 | -0.01 ± 0.03 | 0.838 | 0 ± 0.02 | 0.953 | -0.01 ± 0.01 | 0.371 | **0.02 ± 0.01** | **0.016** | **-0.03 ± 0.01** | **0.043** |

Generalized linear models were used to examine the associations between 16 VOC metabolites and 7 liver biomarkers in the total study population. Since the distributions of both independent variables (VOC metabolites) and dependent variables (7 liver biomarkers) were right skewed, all these variables were log transformed to improve the normality in the models. Models were adjusted for common liver-related confounders: age, sex, race, BMI, diabetes status and alcohol consumption.
The statistical significance was set at the $p$ -value $< 0.05$ . The $\beta$ value shows as 0 if $\beta < 0.01$ . The $p$ -value shows as 0 if $p < 0.001$ . Bold values represent significant associations between VOC metabolites and liver biomarkers. SE: standard errors.

## RESULTS

### Characteristics of the Study Population

After applying the exclusion criteria, the study population consisted of 663 subjects and the demographic characteristics are presented in **Table 1**. In general, the population comprised of a higher number of females (60%) than males (40%), and White persons accounted for 78% of the population. The mean age was 49.3 ± 12.7 years with a range of 25-70 years, and mean body mass index (BMI) was 30.3 ± 6.9 kg/m^2^. When stratified by smoking status, there were more nonsmokers (n=413) than smokers (n=250). A significantly higher number of males were smokers than nonsmokers (45% *vs.* 36% respectively). With regards to race (self-reported), White persons were mostly nonsmokers than smokers (83% *vs.* 69% respectively). In contrast, more Black persons were smokers than nonsmokers (26% *vs.* 12% respectively). Additionally, there were significant differences in age and BMI between nonsmokers and smokers (50 ± 13.2 *vs*. 48.1 ± 11.8 years and 30.9 ± 7.1 *vs.* 29.4 ± 6.5 kg/m^2^ respectively).

### Distribution of exposure biomarkers by smoking status, sex and race

Overall, 16 urinary VOC metabolites (exposure biomarkers) corresponding to 12 parent compounds, were measured in the study population and the percent lower than the limit of detection (% < LOD) for each metabolite is provided in **Table 2**. In general, the metabolites measured were well-detected in the study population, with the exception of 2 metabolites that were not detected in over 30% of the population. These were metabolites of acrylonitrile, namely N-Acetyl-S-(2-cyanoethyl)-L-cysteine (CYMA), and styrene, namely N-Acetyl-S-(1-phenyl-2-hydroxyethyl)-L-cysteine + N-Acetyl-S-(2-phenyl-2-hydroxyethyl)-L-cysteine (PHEMA). The distribution of VOC metabolites within nonsmokers and smokers is provided in **Table 3**. Out of all the VOC metabolites measured, 15 metabolites were significantly higher in smokers *vs.* nonsmokers and these include metabolites of *i)* acrolein, namely N-Acetyl-S-(2-carboxyethyl)-L-cysteine (CEMA) and N-Acetyl-S-(3-hydroxypropyl)-L-cysteine (3HPMA); *ii)* acrylamide, namely N-Acetyl-S-(2-carbamoylethyl)-L-cysteine (AAMA); *iii)* acrylonitrile, namely CYMA; *iv)* benzene, namely trans-Muconic acid (MU); *v)* 1,3-butadiene, namely N-Acetyl-S-(3,4-dihydroxybutyl)-L-cysteine (DHBMA) and N-Acetyl-S-(4-hydroxy-2-buten-1-yl)-L-cysteine (MHBMA3); *vi)* crotonaldehyde, namely N-Acetyl-S-(3-hydroxypropyl-1-methyl)-L-cysteine (HPMMA); *vii)* N,N-dimethylformamide, namely N-Acetyl-S-(N-methylcarbamoyl)-L-cysteine (AMCC); *viii)* ethylbenzene and/or styrene, namely Phenylglyoxylic acid (PGA), PHEMA and Mandelic acid (MA); *ix)* propylene oxide namely N-Acetyl-S-(2-hydroxypropyl)-L-cysteine (2HPMA); and *x)* xylene, namely, 2-Methylhippuric acid (2MHA) and 3-Methylhippuric acid + 4-Methylhippuric acid (3MHA+4MHA). The only metabolite that did not show significantly different levels within smokers and nonsmokers was the toluene metabolite, namely N-Acetyl-S-(benzyl)-L-cysteine (BMA). Metabolites of acrolein, crotonaldehyde and xylene were some of the highest levels detected in smokers, while in nonsmokers, some of the compounds detected at higher levels included metabolites of 1,3-butadine, acrolein, crotonaldehyde and styrene.

When stratified by sex (**Supplemental Table S1A**), there were no significant differences in urinary VOC metabolite levels except for the toluene metabolite (BMA) which was higher in females (n=401) than males (n=262). When stratified by race (**Supplemental Table S1B**), significant differences were observed in VOC metabolite levels where Black persons (n=114) had higher urinary levels of metabolites for acrolein (CEMA), acrylamide (AAMA), acrylonitrile (CYMA), 1,3-butadiene (MHBMA3), toluene (BMA) and xylene (3MHA+4MHA) compared to White persons (n=514).

### Distribution of liver disease biomarkers by smoking status, sex and race

There were no significant differences between smokers’ and nonsmokers’ categories for serum levels of liver disease biomarkers, namely cell death markers (K18 M30, K18 M65, K18 M30/M65), liver enzymes (ALT, AST, ALP) and direct bilirubin (**Supplemental Table S2A**). However, when stratified by sex (**Supplemental Table S2B**), males demonstrated higher levels of serum ALT, AST and direct bilirubin *vs.* females. When stratified by race (**Supplemental Table S2C**), White persons demonstrated higher levels of serum K18 M30, M65, M30/M65 ratio, ALT, and direct bilirubin.

### Relationship between VOC metabolites and liver disease biomarkers in the overall study population

The relationships between urinary VOC metabolites and liver disease biomarkers in the total study population (n=663) are presented in **Table 4** and **Figure 2A**. There were no significant associations between VOC metabolites and K18 M30; however both MU (benzene metabolite) and 2MHA (xylene metabolite) were both negatively associated with K18 M65, while only PGA (ethylbenzene/styrene metabolite) was negatively associated with the calculated M30/M65 ratio. In terms of circulating liver enzymes, there were no significant associations between VOC metabolites and serum ALT and AST. However, 8 VOC metabolites were positively associated with elevated ALP, namely metabolites of acrolein (CEMA and 3HPMA), acrylamide (AAMA), acrylonitrile (CYMA), 1,3-butadine (MHBMA3), crotonaldehyde (HPMMA), styrene (MA) and xylene (3MHA+4MHA). The strongest association was seen for MA where serum ALP increased by 3.4% (95% CI, 1.0% - 5.8%) for every 2-fold increase in urinary MA levels as highlighted by the forest plot (**Figure 3A**). Following MA, metabolites AAMA and CEMA were associated with serum ALP elevation by 3.3% (95% CI, 1.5% - 5.2%) and 2.3% (95% CI, 0.7% - 3.9%), respectively. After adjustment for smoking as a confounder, using either categorical (**Supplemental Table S3 and Figure S1**) or numeric cotinine levels (data not shown), there were no associations between these VOC metabolites and ALP except for the acrylamide metabolite (AAMA). Only the benzene metabolite (MU) was positively associated with serum bilirubin, while 4 metabolites were negatively associated with serum bilirubin and these included metabolites of acrylamide (AAMA), 1,3-butadine (DHBMA), N,N-dimethylformamide (AMCC) and xylene (3MHA+4MHA). Bilirubin was increased by 3.8% (95% CI, 1.3% - 6.3%) for every 2-fold increase in urinary MU levels (**Figure 3A**). After adjustment for smoking as a confounder, using either categorical (**Supplemental Table S4 and Figure S2**) or numeric cotinine levels (data not shown), positive associations were observed for MU and the acrolein metabolite (3HPMA).

**Figure 3.**
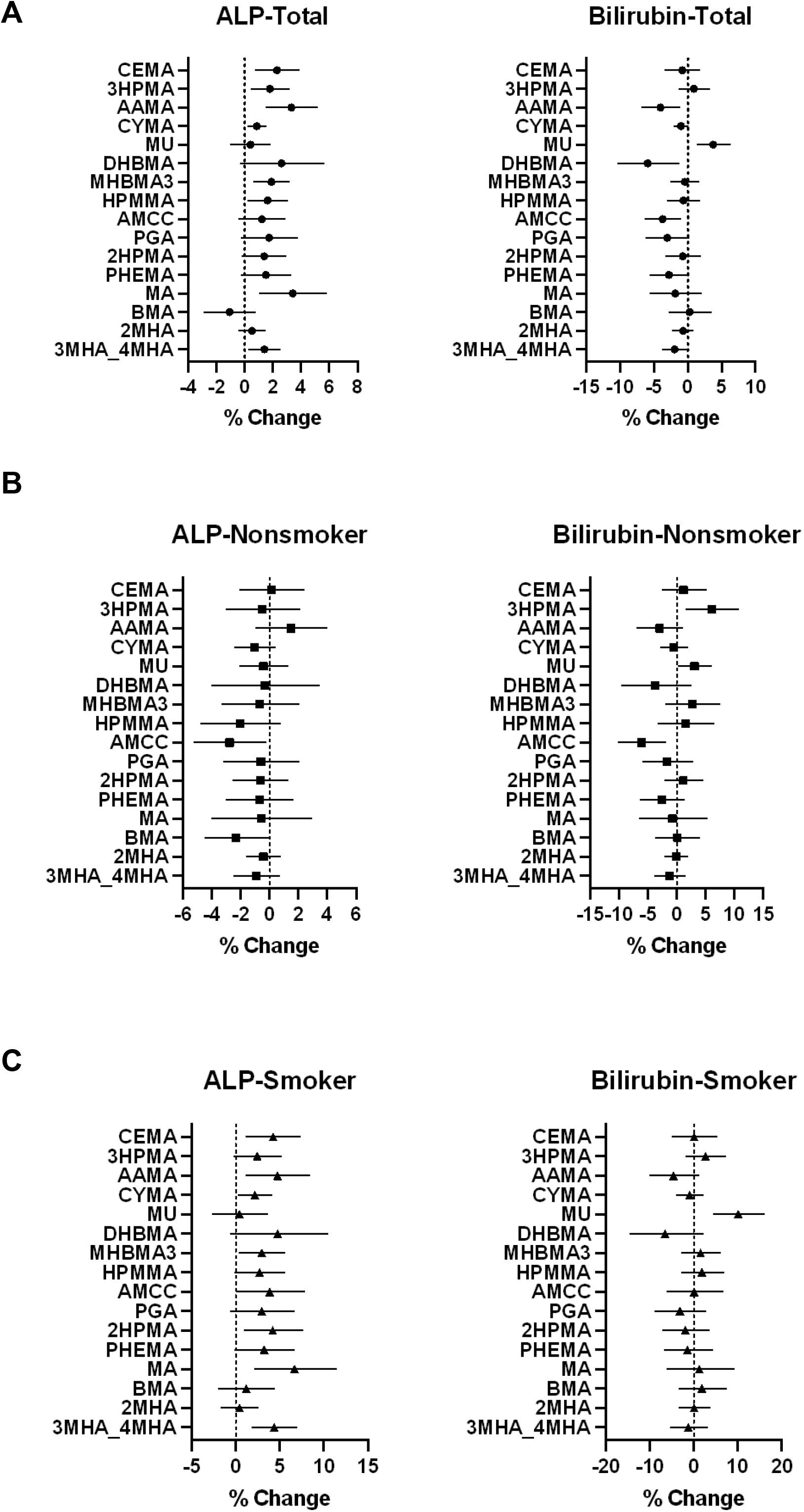
Adjusted associations between urinary levels of 16 VOC metabolites and 2 liver biomarkers (ALP, Bilirubin) in A) the total study population (n = 663), B) nonsmokers (n = 413), and C) smokers (n=250). The forest plots represents % change (95% CI) per 2-fold increase in urinary VOC metabolite levels. Generalized linear models were used to examine the adjusted associations. Models were adjusted for common liver-related confounders: age, sex, race, BMI, diabetes status and alcohol consumption. The statistical significance was set at *p*-value <0.05.

### Relationship between VOC metabolites and liver disease biomarkers within nonsmokers and smokers

The relationships between urinary VOC metabolites and liver disease biomarkers in nonsmokers (n=413) are presented in **Table 5A** and **Figure 2B**. Similar to the total study population, there were no significant associations between VOC metabolites and K18 M30 in nonsmokers. However, PGA (ethylbenzene/styrene metabolite) was positively associated with K18 M65 and negatively associated with the M30/M65 ratio in nonsmokers; while 4 VOC metabolites were negatively associated with K18 M65, and these included metabolites of acrolein (3HPMA), N,N-dimethylformamide (AMCC), styrene (PHEMA) and xylene (2MHA). Additionally, 2MHA (xylene metabolite) was also positively associated with M30/M65. Negative associations were observed for the N,N-dimethylformamide metabolite (AMCC) with AST, ALP and bilirubin in nonsmokers, while the toluene metabolite (BMA) was also negatively associated with ALP. Interestingly, both metabolites of acrolein (3HPMA) and benzene (MU) were positively associated with serum bilirubin in nonsmokers. Indeed, bilirubin increased by 6.0% (95% CI, 1.5% - 10.8%) and 3.1% (95% CI, 0.1% - 6.0%) per 2-fold increase of urinary 3HPMA and MU levels, respectively, in nonsmokers (**Figure 3B**)

**Table 5A.** Relationships between the 16 VOC metabolites and 7 liver biomarkers in nonsmokers (n=413).

| VOC metabolites | Keratin 18 (n = 413) |  |  |  |  |  | Enzymes and Bilirubin (n = 413) |  |  |  |  |  |  |  |
| --- | --- | --- | --- | --- | --- | --- | --- | --- | --- | --- | --- | --- | --- | --- |
|  | M30 |  | M65 |  | M30/M65 |  | ALT |  | AST |  | ALP |  | Bilirubin |  |
| | $\beta \pm SE$ | p-value | $\beta \pm SE$ | p-value | $\beta \pm SE$ | p-value | $\beta \pm SE$ | p-value | $\beta \pm SE$ | p-value | $\beta \pm SE$ | p-value | $\beta \pm SE$ | p-value |
| CEMA | 0 ± 0.06 | 0.953 | -0.04 ± 0.04 | 0.31 | 0.04 ± 0.06 | 0.462 | 0.06 ± 0.04 | 0.127 | 0.01 ± 0.03 | 0.723 | 0 ± 0.02 | 0.902 | 0.02 ± 0.03 | 0.557 |
| 3HPMA | 0.01 ± 0.07 | 0.843 | **-0.1 ± 0.04** | **0.022** | 0.11 ± 0.06 | 0.084 | -0.01 ± 0.04 | 0.779 | 0.01 ± 0.03 | 0.714 | -0.01 ± 0.02 | 0.703 | **0.08 ± 0.03** | **0.008** |
| AAMA | 0.04 ± 0.07 | 0.529 | -0.04 ± 0.04 | 0.305 | 0.08 ± 0.06 | 0.169 | -0.01 ± 0.04 | 0.726 | -0.03 ± 0.03 | 0.224 | 0.02 ± 0.02 | 0.238 | -0.04 ± 0.03 | 0.147 |
| CYMA | -0.01 ± 0.04 | 0.898 | -0.04 ± 0.02 | 0.105 | 0.03 ± 0.04 | 0.354 | 0 ± 0.02 | 0.974 | -0.02 ± 0.02 | 0.289 | -0.01 ± 0.01 | 0.162 | -0.01 ± 0.02 | 0.695 |
| MU | -0.04 ± 0.05 | 0.439 | -0.05 ± 0.03 | 0.099 | 0.01 ± 0.04 | 0.815 | -0.01 ± 0.03 | 0.809 | -0.01 ± 0.02 | 0.606 | -0.01 ± 0.01 | 0.625 | **0.04 ± 0.02** | **0.04** |
| DHBMA | -0.01 ± 0.1 | 0.911 | 0.01 ± 0.06 | 0.856 | -0.02 ± 0.09 | 0.808 | -0.05 ± 0.06 | 0.399 | -0.04 ± 0.04 | 0.302 | -0.01 ± 0.03 | 0.854 | -0.06 ± 0.05 | 0.24 |
| MHBMA3 | -0.06 ± 0.07 | 0.393 | -0.03 ± 0.04 | 0.499 | -0.03 ± 0.07 | 0.621 | 0.02 ± 0.05 | 0.67 | 0.02 ± 0.03 | 0.582 | -0.01 ± 0.02 | 0.621 | 0.04 ± 0.03 | 0.271 |
| HPMMA | 0.06 ± 0.08 | 0.451 | 0 ± 0.05 | 0.968 | 0.06 ± 0.07 | 0.421 | 0.06 ± 0.05 | 0.192 | 0.06 ± 0.03 | 0.061 | -0.03 ± 0.02 | 0.151 | 0.02 ± 0.04 | 0.549 |
| AMCC | -0.13 ± 0.07 | 0.063 | **-0.09 ± 0.04** | **0.028** | -0.04 ± 0.06 | 0.548 | 0.02 ± 0.04 | 0.641 | **-0.07 ± 0.03** | **0.015** | **-0.04 ± 0.02** | **0.034** | **-0.09 ± 0.03** | **0.005** |
| PGA | -0.08 ± 0.07 | 0.265 | **0.09 ± 0.04** | **0.039** | **-0.17 ± 0.07** | **0.009** | 0.02 ± 0.05 | 0.598 | -0.04 ± 0.03 | 0.237 | -0.01 ± 0.02 | 0.652 | -0.02 ± 0.03 | 0.457 |
| 2HPMA | -0.02 ± 0.05 | 0.667 | -0.05 ± 0.03 | 0.15 | 0.02 ± 0.05 | 0.635 | -0.02 ± 0.03 | 0.487 | -0.02 ± 0.02 | 0.27 | -0.01 ± 0.01 | 0.523 | 0.02 ± 0.02 | 0.519 |
| PHEMA | -0.09 ± 0.06 | 0.142 | **-0.09 ± 0.04** | **0.027** | -0.01 ± 0.06 | 0.875 | -0.05 ± 0.04 | 0.2 | -0.03 ± 0.03 | 0.289 | -0.01 ± 0.02 | 0.557 | -0.04 ± 0.03 | 0.191 |
| MA | -0.05 ± 0.1 | 0.607 | -0.04 ± 0.06 | 0.52 | -0.01 ± 0.09 | 0.887 | 0 ± 0.06 | 0.978 | -0.07 ± 0.04 | 0.079 | -0.01 ± 0.03 | 0.74 | -0.01 ± 0.04 | 0.799 |
| BMA | -0.05 ± 0.06 | 0.425 | -0.04 ± 0.04 | 0.293 | -0.01 ± 0.06 | 0.853 | -0.02 ± 0.04 | 0.582 | -0.02 ± 0.03 | 0.352 | **-0.03 ± 0.02** | **0.043** | 0 ± 0.03 | 0.968 |
| 2MHA | 0.01 ± 0.03 | 0.73 | **-0.05 ± 0.02** | **0.006** | **0.07 ± 0.03** | **0.029** | 0 ± 0.02 | 0.944 | -0.01 ± 0.01 | 0.532 | -0.01 ± 0.01 | 0.486 | 0 ± 0.02 | 0.918 |
| 3MHA+4MHA | 0 ± 0.04 | 0.962 | -0.01 ± 0.03 | 0.843 | 0 ± 0.04 | 0.937 | -0.01 ± 0.03 | 0.673 | -0.02 ± 0.02 | 0.192 | -0.01 ± 0.01 | 0.271 | -0.02 ± 0.02 | 0.359 |

Generalized linear models were used to examine the associations between 16 VOC metabolites and 7 liver biomarkers in the total study population. Since the distributions of both independent variables (VOC metabolites) and dependent variables (7 liver biomarkers) were right skewed, all these variables were log transformed to improve the normality in the models. Models were adjusted for common liver-related confounders: age, sex, race, BMI, diabetes status and alcohol consumption.
The statistical significance was set at the $p$ -value $< 0.05$ . The $\beta$ value shows as 0 if $\beta < 0.01$ . The $p$ -value shows as 0 if $p < 0.001$ . Bold values represent significant associations between VOC metabolites and liver biomarkers. SE: standard errors.

The relationship between urinary VOC metabolites and liver disease biomarkers in smokers (n=250) are presented in **Table 5B** and **Figure 2C**. There were no significant associations between any of the urinary VOC metabolites and serum K18 M30 and M30/M65 ratio in smokers. However, the benzene metabolite (MU) was negatively associated with K18 M65 in smokers, an observation that was seen in the total population as well. Regarding circulating liver enzymes, the styrene metabolite (MA) was positively associated with both ALT and AST. In addition, HPMMA (crotonaldehyde metabolite) was positively associated with serum AST. Furthermore, 8 VOC metabolites were positively associated with serum ALP, including metabolites of acrolein (CEMA), acrylonitrile (AAMA), acrylamide (CYMA), 1,3-butadine (MHBMA3), N,N-dimethylformamide (AMCC), propylene oxide (2HPMA), styrene (MA) and xylene (3MHA+4MHA). The strongest association was seen for MA where serum ALP increased by 6.7% (95% CI, 2.1% - 11.5%) for every 2-fold increase in urinary MA levels in smokers followed by AAMA for a 4.7% increase (95% CI, 1.1% - 8.5%) and 3MHA+4MHA for a 4.4% increase (95% CI, 1.8% - 7.0%) (**Figure 3C**). Most of these metabolites also showed significant associations in the total population. Furthermore, similar to both the total population and nonsmokers’ category, the benzene metabolite (MU) also showed a positive association with serum bilirubin in smokers. Bilirubin increased by 10.1% (95% CI, 4.4% - 16.1%) for every 2-fold increase in urinary MU levels (**Figure 3C**).

**Table 5B.** Relationships between the 16 VOC metabolites and 7 liver biomarkers in smokers (n=250).

| VOC metabolites | Keratin 18 (n = 250) |  |  |  |  |  | Enzymes and Bilirubin (n = 250) |  |  |  |  |  |  |  |
| --- | --- | --- | --- | --- | --- | --- | --- | --- | --- | --- | --- | --- | --- | --- |
|  | M30 |  | M65 |  | M30/M65 |  | ALT |  | AST |  | ALP |  | Bilirubin |  |
| | $\beta \pm SE$ | p-value | $\beta \pm SE$ | p-value | $\beta \pm SE$ | p-value | $\beta \pm SE$ | p-value | $\beta \pm SE$ | p-value | $\beta \pm SE$ | p-value | $\beta \pm SE$ | p-value |
| CEMA | -0.04 ± 0.09 | 0.605 | 0 ± 0.05 | 0.977 | -0.04 ± 0.07 | 0.568 | 0.04 ± 0.05 | 0.443 | 0.02 ± 0.04 | 0.679 | **0.06 ± 0.02** | **0.007** | 0 ± 0.04 | 0.997 |
| 3HPMA | 0.06 ± 0.08 | 0.411 | 0.01 ± 0.04 | 0.752 | 0.05 ± 0.07 | 0.463 | 0.05 ± 0.05 | 0.273 | 0.05 ± 0.03 | 0.087 | 0.03 ± 0.02 | 0.082 | 0.04 ± 0.03 | 0.269 |
| AAMA | 0.01 ± 0.1 | 0.916 | -0.03 ± 0.06 | 0.64 | 0.04 ± 0.09 | 0.674 | -0.08 ± 0.06 | 0.172 | -0.08 ± 0.04 | 0.063 | **0.07 ± 0.03** | **0.01** | -0.07 ± 0.04 | 0.12 |
| CYMA | 0.05 ± 0.05 | 0.348 | 0.03 ± 0.03 | 0.285 | 0.02 ± 0.05 | 0.701 | 0.02 ± 0.03 | 0.573 | 0.04 ± 0.02 | 0.076 | **0.03 ± 0.01** | **0.025** | -0.01 ± 0.02 | 0.535 |
| MU | -0.12 ± 0.09 | 0.169 | **-0.14 ± 0.05** | **0.005** | 0.02 ± 0.08 | 0.826 | -0.04 ± 0.05 | 0.491 | 0 ± 0.04 | 0.918 | 0.01 ± 0.02 | 0.812 | **0.14 ± 0.04** | **0** |
| DHBMA | -0.09 ± 0.15 | 0.569 | 0.06 ± 0.08 | 0.477 | -0.15 ± 0.13 | 0.269 | -0.02 ± 0.09 | 0.831 | 0.02 ± 0.06 | 0.732 | 0.07 ± 0.04 | 0.089 | -0.1 ± 0.07 | 0.141 |
| MHBMA3 | -0.02 ± 0.07 | 0.764 | 0.01 ± 0.04 | 0.836 | -0.03 ± 0.06 | 0.635 | 0.03 ± 0.04 | 0.468 | 0.04 ± 0.03 | 0.237 | **0.04 ± 0.02** | **0.028** | 0.02 ± 0.03 | 0.505 |
| HPMMA | 0.04 ± 0.08 | 0.582 | 0.02 ± 0.04 | 0.681 | 0.03 ± 0.07 | 0.716 | 0.07 ± 0.05 | 0.125 | **0.08 ± 0.03** | **0.016** | 0.04 ± 0.02 | 0.066 | 0.03 ± 0.04 | 0.461 |
| AMCC | 0.05 ± 0.11 | 0.613 | 0.04 ± 0.06 | 0.471 | 0.01 ± 0.09 | 0.909 | 0.08 ± 0.06 | 0.228 | 0.07 ± 0.04 | 0.096 | **0.05 ± 0.03** | **0.049** | 0 ± 0.05 | 0.996 |
| PGA | -0.12 ± 0.1 | 0.241 | -0.02 ± 0.06 | 0.75 | -0.1 ± 0.09 | 0.258 | 0.01 ± 0.06 | 0.884 | 0 ± 0.04 | 0.943 | 0.04 ± 0.03 | 0.112 | -0.05 ± 0.04 | 0.295 |
| 2HPMA | 0.01 ± 0.09 | 0.896 | 0 ± 0.05 | 0.954 | 0.02 ± 0.08 | 0.853 | -0.01 ± 0.06 | 0.854 | -0.02 ± 0.04 | 0.625 | **0.06 ± 0.02** | **0.013** | -0.03 ± 0.04 | 0.482 |
| PHEMA | -0.09 ± 0.09 | 0.312 | -0.02 ± 0.05 | 0.72 | -0.08 ± 0.08 | 0.357 | -0.01 ± 0.06 | 0.874 | 0.05 ± 0.04 | 0.236 | 0.05 ± 0.02 | 0.06 | -0.02 ± 0.04 | 0.611 |
| MA | 0.04 ± 0.13 | 0.761 | 0.04 ± 0.07 | 0.563 | 0 ± 0.11 | 0.981 | **0.16 ± 0.08** | **0.039** | **0.11 ± 0.05** | **0.041** | **0.09 ± 0.03** | **0.004** | 0.02 ± 0.06 | 0.756 |
| BMA | 0.04 ± 0.09 | 0.682 | 0.01 ± 0.05 | 0.833 | 0.03 ± 0.08 | 0.741 | 0.05 ± 0.05 | 0.361 | 0.03 ± 0.04 | 0.392 | 0.02 ± 0.02 | 0.474 | 0.03 ± 0.04 | 0.526 |
| 2MHA | -0.02 ± 0.06 | 0.737 | -0.03 ± 0.03 | 0.335 | 0.01 ± 0.05 | 0.815 | 0.02 ± 0.04 | 0.655 | 0.01 ± 0.03 | 0.631 | 0.01 ± 0.02 | 0.714 | 0 ± 0.03 | 1 |
| 3MHA+4MHA | -0.02 ± 0.07 | 0.774 | 0 ± 0.04 | 0.999 | -0.02 ± 0.06 | 0.744 | 0.03 ± 0.04 | 0.561 | 0.01 ± 0.03 | 0.726 | **0.06 ± 0.02** | **0.001** | -0.02 ± 0.03 | 0.573 |

Generalized linear models were used to examine the associations between 16 VOC metabolites and 7 liver biomarkers in the total study population. Since the distributions of both independent variables (VOC metabolites) and dependent variables (7 liver biomarkers) were right skewed, all these variables were log transformed to improve the normality in the models. Models were adjusted for common liver-related confounders: age, sex, race, BMI, diabetes status and alcohol consumption.
The statistical significance was set at the $p$ -value $< 0.05$ . The $\beta$ value shows as 0 if $\beta < 0.01$ . The $p$ -value shows as 0 if $p < 0.001$ . Bold values represent significant associations between VOC metabolites and liver biomarkers. SE: standard errors.

## DISCUSSION

Numerous epidemiologic studies have demonstrated that exposures to VOCs were associated with various forms of liver injury and dysfunction ranging from elevated liver enzyme levels to steatohepatitis and liver cancer. For instance, it is well-documented that exposure to vinyl chloride, a historic occupational VOC, was associated with steatohepatitis in rubber plant workers, with some workers exhibiting hemangiosarcoma, a rare form of cancer affecting liver sinusoidal cells (Cave, et al., 2010; Fedeli *et al*., 2019). Other studies focusing on petroleum and auto-industry workers demonstrated that workers who were occupationally exposed to VOCs such as gasoline and other aromatic hydrocarbons, had higher mean levels of liver test results (ALT, AST and ALP) as well as total and conjugated bilirubin, when compared to non-worker controls (Asefaw *et al*., 2020; Egbuonu and Nkwazema, 2015; Mohammadi *et al*., 2010). Similar associations have also been observed in non-occupational exposure settings. For example, in a study conducted using the National Health and Nutrition Examination Survey (NHANES 1999-2000) database, personal exposures to VOCs including benzene, ethylbenzene, toluene, and xylene were positively correlated with serum ALP and total bilirubin levels (Liu *et al*., 2009). Furthermore, in a cross-sectional study examining environmental VOC exposures and metabolic parameters in the Canadian population, ALP and AST were associated with higher blood concentrations of xylene, styrene and toluene (Cakmak *et al*., 2020). However, such studies looking at residential or environmental VOC exposures and liver disease, are still scarce.

In the current study, we examined the association between 16 urinary VOC metabolites and liver disease biomarkers, namely biomarkers for liver cell death (K18 M30 and M65) and liver chemistry tests (ALT, AST, ALP and bilirubin). Smokers had higher urinary VOC metabolite levels than nonsmokers, which is in concordance with previous reports (Chambers *et al*., 2011; Nazaroff and Singer, 2004). Out of all the metabolites measured, 8 VOC metabolites corresponding to acrolein, acrylonitrile, acrylamide, 1,3-butadiene, crotonaldehyde, styrene and xylene were positively associated with serum ALP in the overall population, which is a major finding of the study. In smokers, these same VOC metabolites, with the addition N,N dimethylformamide and propylene oxide metabolites, and the exception of crotonaldehyde metabolite, were positively associated with ALP, implicating that tobacco smoke is a source of VOC exposure, resulting in the observed higher VOC metabolite body burdens in smokers. Based on these results, it could be argued that smokers may be more prone to liver injury and dysfunction, as smoking has been proposed to contribute to early liver disease onset and advanced liver fibrosis (Jung *et al*., 2019; Mantaka *et al*., 2018; Okamoto *et al*., 2018). After adjusting for smoking as a confounder, only the acrylamide metabolite (AAMA) demonstrated a positive association with serum ALP, suggesting that the AAMA effect potentially came from non-tobacco smoke sources such as heated foods (Semla *et al*., 2017). Notably, serum ALP upregulation often occurs in the presence of bile duct obstruction, and tests for ALP along with bilirubin can be used to determine liver disease of a cholestatic pattern (Kwo *et al*., 2017; Lala *et al*., 2021). Moreover, because smokers have been reported to be at greater risk for cholestatic diseases (Huang *et al*., 2017; Smyk *et al*., 2012), it can be postulated that the VOC associations with ALP in smokers, in the current study, could possibly be related to cholestatic injury. Another compelling observation in the study was the consistent positive association between the benzene metabolite (MU) and bilirubin in both smokers and nonsmokers, although the association was stronger in smokers (10.1% *vs.* 3.1% increase in bilirubin for every 2-fold increase in urinary VOC metabolite level). After adjusting for smoking as a confounder, the positive association between MU and bilirubin still prevailed. In addition, the acrolein metabolite (3HPMA) was also positively associated with bilirubin but only in nonsmokers, highlighting that some VOCs including benzene may contribute to altered liver disease biomarkers (Werder, et al., 2020), in the absence of smoking.

There were no positive associations between the measured VOC metabolites and K18 biomarkers, except for the ethylbenzene/styrene metabolite, implicating that these VOCs did not impact hepatocyte death. The lack of statistically significant associations between the measured VOC metabolites and K18 biomarkers could possibly be due to the fact that these are residential exposures, and K18 biomarkers are more responsive to occupational, higher-dose exposures, or in populations with a known history of exposures (Cave, et al., 2011; Clair, et al., 2018; Werder, et al., 2020). Nonetheless, the associations observed for ALP and bilirubin reinforces the negative role of VOCs, coupled with lifestyle factors such as smoking, on liver health and function. Importantly, sex and race also influenced both exposure and liver disease biomarker levels in the study population. While most males tended to be smokers consistent with previous reports (Agaku *et al*., 2020), there was no significant sex differences in urinary VOC metabolite levels except for the toluene metabolite which was higher in females. Toluene is a major contaminant emitted from cosmetic products (Zhou *et al*., 2016), and may have led to higher levels in females. Nonetheless, females displayed lower levels of liver disease biomarkers namely ALT, AST and bilirubin than males, consistent with previous findings (Mera *et al*., 2008). With regards to race, more smokers tended to be Black persons than White persons, and this was also reflected in exposure biomarker levels with Black persons having higher urinary metabolite levels for acrolein, acrylonitrile and other tobacco smoke products. Previous studies examining disparities with smoking reported that similar percentages of White and Black persons who smoke cigarettes while cigar use was more prevalent in Black persons (Agaku, et al., 2020; Weinberger *et al*., 2020). These findings advocate for the importance of considering sex and race as potential mediators, when assessing risk of residential exposures to chemicals such as VOCs that are ubiquitous, and likely regulated by lifestyle factors and socio-economic status.

The potential mechanisms driving the observed hepatotoxicity associated with VOC exposures have been investigated in model systems. Rodent studies evaluating VOC liver effects at varying doses demonstrated upregulated serum ALT, AST and ALP, indicative of liver injury, accompanied by immune-modulating responses and hepatic metabolic disruption (Abplanalp *et al*., 2019; Dere and Ari, 2009; Lang *et al*., 2018; Wahlang *et al*., 2020; Wang *et al*., 2016). VOCs also appear to exacerbate liver disease endpoints, such as NAFLD and overall energy homeostasis, in conjunction with additional factors such as high caloric diets (Lang, et al., 2018; Wahlang, et al., 2020). As xenobiotics, VOCs undergo biotransformation in the liver and can form reactive intermediates during the detoxification process. Most VOCs convert to reactive electrophiles in the liver and can bind to cellular macromolecules to form adducts (Rusyn *et al*., 2021), which is a key characteristic for vinyl chloride-induced toxicity. Other mechanism(s) of VOC toxicity that have been postulated include induction of oxidative and endoplasmic reticulum (ER) stress, mitochondrial dysfunction, activation of immune-mediated responses and cell death, and metabolic disruption through xenobiotic and endobiotic receptor activation (Lang and Beier, 2018; Rusyn, et al., 2021; Wahlang, et al., 2019). However, mechanistic studies pertinent to the VOCs discussed in the current study, and their causal roles in liver disease initiation and progression, are still lacking and require future investigations.

A major strength of the current study is that it is one of the first few to evaluate liver injury outcomes in the context of residential, low dose exposures to VOCs, while also accounting the influence of smoking; it is however not without limitations. A major concern in the current study is the source of serum ALP that was measured, since ALP is not liver-specific, and could also originate from other sources such as bone and kidney. However, because the liver is the largest body organ, it does contribute to at least 80% of serum ALP (Lowe *et al*., 2021). Another limitation is that while the study focused on VOC exposures in community residents, it did not adjust for occupation and the likelihood that some residents may be exposed to VOCs at work rather than within their home environment. In addition, another potential limitation of the study is that it fails to address reverse causality, and the lack of a longitudinal set-up to determine if liver disease impacted VOC body burdens or if VOC levels influence liver disease biomarkers.

Nevertheless, these limitations will be addressed in our future studies using a longitudinal cohort to address VOC exposure risks and cardio-metabolic health, and if lifestyle factors such as smoking are potential mediators of VOC associations with liver injury markers. Elevation of gamma-glutamyl transferase (GGT) will also be measured, in the proposed cohort, to confirm if the ALP effect is liver-specific (Kwo, et al., 2017). Importantly, because we are investigating multiple VOCs simultaneously, implementing multi-pollutant models will be another approach to analyze VOC associations with liver disease biomarkers.

In conclusion, the results from our current study demonstrated that residential exposures to VOCs were positively associated with liver injury markers, particularly ALP, in the overall population. These associations were also observed in smokers, and we postulate that smoking may have attributed to both higher VOC levels in smokers and the observed associations. Racial and sex differences were also noted in relation to exposure and disease biomarker levels. The findings from this cross-sectional study provide some insight into how residential, low dose VOC exposures can influence liver injury markers, and the potential impact of smoking and tobacco use in the context of VOCs in NAFLD and TASH warrant further investigations. Such studies will allow for better risk assessment in terms of daily VOC exposures, and how lifestyle factors such as smoking can modify risk. Overall, the current findings provide substantial evidence that residential VOCs exposures may potential affect liver health and function, and in-depth, translational, mechanistic studies are needed to confirm if the observed associations are causal.

## Data Availability

N/A

## FUNDING SUPPORT

This research was supported, in part, by the National Institute of Environmental Health Sciences (P42ES023716, P30ES030283, R35ES028373, R01ES032189, R01ES029846 and R21ES031510); the National Heart Blood and Lung Institute (U54HL120163); the National Institute of General Medical Sciences (P20GM113226); and the National Institute on Alcohol Abuse and Alcoholism (P50AA024337).

## CONFLICT OF INTEREST

The authors declare that they have no conflict of interest.

## ACKNOWLEDGEMENTS

The authors would like to thank all the partners, members and volunteers involved in the HEAL Study.

---

**Supplemental Table S1A:** Distribution of VOC metabolites within males and females.

| VOC metabolites | Total (n = 663) |  |  | Males (n = 262) |  |  | Females (n = 401) |  |  | p-value |
| --- | --- | --- | --- | --- | --- | --- | --- | --- | --- | --- |
| | Mean ± SD | Median ± IQR | Min - Max | Mean ± SD | Median ± IQR | Min - Max | Mean ± SD | Median ± IQR | Min - Max | |
| CEMA | 193.4 ± 219 | 117.8 ± 163.6 | 3.8 - 1981.2 | 188.7 ± 190.4 | 114.6 ± 186.4 | 3.8 - 1114.3 | 196.6 ± 236 | 120.4 ± 150.8 | 5.6 - 1981.2 | 0.817 |
| 3HPMA | 872.5 ± 1217.5 | 367.2 ± 833.7 | 46.3 - 9597.4 | 853.7 ± 1097.3 | 381.9 ± 878.4 | 59.5 - 7593.8 | 884.8 ± 1291.3 | 360.7 ± 819.3 | 46.3 - 9597.4 | 0.542 |
| AAMA | 109.1 ± 122.9 | 73.9 ± 92.9 | 4.7 - 1300.9 | 115.9 ± 128.3 | 80.8 ± 93.8 | 4.7 - 1231.2 | 104.6 ± 119.3 | 69.7 ± 85.8 | 8.4 - 1300.9 | 0.198 |
| CYMA | 59.5 ± 108.2 | 3.6 ± 83.7 | 0.2 - 883.3 | 66.6 ± 110.8 | 4.1 ± 102.5 | 0.3 - 798 | 54.9 ± 106.3 | 3.5 ± 72.6 | 0.2 - 883.3 | 0.217 |
| MU | 149.7 ± 199.3 | 86.6 ± 141.7 | 6 - 1827 | 132.6 ± 185.2 | 78 ± 117.7 | 6 - 1503.1 | 160.9 ± 207.5 | 89.2 ± 154.3 | 7.8 - 1827 | 0.074 |
| DHBMA | 403.2 ± 213 | 361.5 ± 246.6 | 50.2 - 1947.3 | 406 ± 203.7 | 365.7 ± 262.4 | 62.1 - 1315.7 | 401.5 ± 219 | 358.7 ± 233.6 | 50.2 - 1947.3 | 0.697 |
| MHBMA3 | 28.1 ± 39.3 | 9.8 ± 29.7 | 0.8 - 293.9 | 28.2 ± 39.2 | 9.4 ± 33.3 | 1 - 293.9 | 28 ± 39.5 | 9.9 ± 25.4 | 0.8 - 240.5 | 0.917 |
| HPMMA | 760.1 ± 947.1 | 319.7 ± 781.5 | 13.5 - 5644.9 | 766.4 ± 924 | 317.8 ± 895.3 | 13.5 - 4981.5 | 756 ± 963.1 | 319.7 ± 678.2 | 50.5 - 5644.9 | 0.773 |
| AMCC | 295.5 ± 290.9 | 195.1 ± 271.7 | 2.6 - 2547.1 | 273.8 ± 258.9 | 187.4 ± 266.2 | 12.6 - 1527.4 | 309.7 ± 309.5 | 198.5 ± 272.4 | 2.6 - 2547.1 | 0.100 |
| PGA | 300.1 ± 211.4 | 248.3 ± 216.3 | 3.4 - 1700.4 | 295.4 ± 212 | 244.2 ± 212.5 | 9.8 - 1700.4 | 303.1 ± 211.2 | 253.5 ± 216.5 | 3.4 - 1592.4 | 0.697 |
| 2HPMA | 68.9 ± 109.5 | 37.3 ± 54.5 | 3.4 - 1044.7 | 72.9 ± 125.7 | 38 ± 54.1 | 3.4 - 1044.7 | 66.3 ± 97.4 | 37 ± 57.6 | 4.3 - 1020.8 | 0.833 |
| PHEMA | 2.8 ± 4 | 1.8 ± 2.1 | 0.2 - 52.6 | 3.1 ± 4.7 | 1.9 ± 2.4 | 0.2 - 52.6 | 2.6 ± 3.3 | 1.8 ± 1.9 | 0.2 - 38.9 | 0.794 |
| MA | 321.8 ± 249.4 | 247.9 ± 228 | 20.9 - 2681.5 | 317.3 ± 243.2 | 233 ± 237.6 | 43.6 - 1506.1 | 324.8 ± 253.7 | 250.5 ± 222 | 20.9 - 2681.5 | 0.386 |
| BMA | 11.5 ± 12.7 | 7.4 ± 8.8 | 0.1 - 106.9 | **10.7 ± 11.7** | 6.7 ± 8.5 | 0.1 - 78.6 | **12 ± 13.3** | 7.9 ± 8.5 | 0.9 - 106.9 | **0.013** |
| 2MHA | 56.1 ± 94.3 | 24.1 ± 55.4 | 0.2 - 1058.9 | 52.5 ± 67.8 | 26.1 ± 55.1 | 0.3 - 532.6 | 58.5 ± 108.2 | 23.7 ± 56.2 | 0.2 - 1058.9 | 0.092 |
| 3MHA+4MHA | 425.3 ± 669 | 174.9 ± 501.2 | 3.4 - 9253.9 | 462.7 ± 800.9 | 193.1 ± 533.6 | 3.4 - 9253.9 | 400.8 ± 566.2 | 166.7 ± 483.5 | 3.4 - 5499.1 | 0.221 |

All the VOC metabolites were normalized by the urinary creatinine. Units: $\mu\text{g/g}$ creatinine.
SD: standard deviation, IQR: the interquartile range, min: minimum, max: maximum
Statistical tests for the comparison between nonsmokers and smokers: the t-test of log-transformed VOC metabolites. Bold values represent significant differences between these two subgroups.

**Supplemental Table S1B:** Distribution of VOC metabolites within White and Black persons.

| VOC metabolites | Total (n = 663) |  |  | White (n = 514) |  |  | Black (n = 114) |  |  | p-value |
| --- | --- | --- | --- | --- | --- | --- | --- | --- | --- | --- |
| | Mean ± SD | Median ± IQR | Min - Max | Mean ± SD | Median ± IQR | Min - Max | Mean ± SD | Median ± IQR | Min - Max | |
| CEMA | 193.4 ± 219 | 117.8 ± 163.6 | 3.8 - 1981.2 | **190.8 ± 226.3** | 115.1 ± 155.4 | 3.8 - 1981.2 | **214.7 ± 179.4** | 151.3 ± 187.1 | 25.9 - 975.8 | **0.001** |
| 3HPMA | 872.5 ± 1217.5 | 367.2 ± 833.7 | 46.3 - 9597.4 | 871.9 ± 1270.4 | 354.4 ± 813.7 | 46.3 - 9597.4 | 884.6 ± 1030.2 | 519.6 ± 866.9 | 57.1 - 5167.4 | 0.183 |
| AAMA | 109.1 ± 122.9 | 73.9 ± 92.9 | 4.7 - 1300.9 | **104 ± 125.5** | 68.9 ± 85.6 | 4.7 - 1300.9 | **127.3 ± 89.5** | 108.7 ± 108.4 | 9.5 - 484.2 | **<.0001** |
| CYMA | 59.5 ± 108.2 | 3.6 ± 83.7 | 0.2 - 883.3 | **54.5 ± 109.4** | 2.7 ± 64.5 | 0.2 - 883.3 | **83.2 ± 104.6** | 45.2 ± 109.9 | 0.2 - 550.1 | **<.0001** |
| MU | 149.7 ± 199.3 | 86.6 ± 141.7 | 6 - 1827 | 156.3 ± 202.8 | 92 ± 149 | 6 - 1827 | 126.2 ± 176.8 | 77 ± 114.9 | 7.8 - 1231 | 0.096 |
| DHBMA | 403.2 ± 213 | 361.5 ± 246.6 | 50.2 - 1947.3 | 413.3 ± 220.5 | 367.6 ± 263.6 | 50.2 - 1947.3 | 381.2 ± 188.6 | 336.5 ± 215.3 | 95.6 - 1287.1 | 0.235 |
| MHBMA3 | 28.1 ± 39.3 | 9.8 ± 29.7 | 0.8 - 293.9 | **28.2 ± 40.9** | 9.2 ± 25.5 | 0.8 - 293.9 | **29.6 ± 34.3** | 13.8 ± 35.1 | 1.6 - 240.5 | **0.040** |
| HPMMA | 760.1 ± 947.1 | 319.7 ± 781.5 | 13.5 - 5644.9 | 760.7 ± 981.3 | 299.9 ± 737.7 | 13.5 - 5644.9 | 800.8 ± 845.4 | 465.6 ± 822 | 50.5 - 3887.6 | 0.073 |
| AMCC | 295.5 ± 290.9 | 195.1 ± 271.7 | 2.6 - 2547.1 | 303.4 ± 303.3 | 193.2 ± 273.7 | 12.6 - 2547.1 | 281.4 ± 245.5 | 202.8 ± 265.9 | 11.8 - 1206.9 | 0.565 |
| PGA | 300.1 ± 211.4 | 248.3 ± 216.3 | 3.4 - 1700.4 | 297.7 ± 211.6 | 244.3 ± 210.8 | 3.4 - 1592.4 | 322.7 ± 224.5 | 289.2 ± 251.6 | 17.2 - 1700.4 | 0.197 |
| 2HPMA | 68.9 ± 109.5 | 37.3 ± 54.5 | 3.4 - 1044.7 | 69.7 ± 115.4 | 35.7 ± 55.2 | 3.4 - 1044.7 | 57 ± 42.6 | 42.2 ± 50.4 | 9 - 264.6 | 0.106 |
| PHEMA | 2.8 ± 4 | 1.8 ± 2.1 | 0.2 - 52.6 | 2.7 ± 4 | 1.8 ± 2 | 0.2 - 52.6 | 3.3 ± 3.9 | 2.4 ± 2.5 | 0.2 - 28.1 | 0.051 |
| MA | 321.8 ± 249.4 | 247.9 ± 228 | 20.9 - 2681.5 | 325.3 ± 256.7 | 247.2 ± 231.8 | 43.6 - 2681.5 | 328 ± 236.9 | 262.2 ± 257.6 | 20.9 - 1506.1 | 0.992 |
| BMA | 11.5 ± 12.7 | 7.4 ± 8.8 | 0.1 - 106.9 | **11.1 ± 12.3** | 7.1 ± 8.3 | 0.1 - 106.9 | **13.7 ± 14.9** | 8.9 ± 9.9 | 1.1 - 93.1 | **0.018** |
| 2MHA | 56.1 ± 94.3 | 24.1 ± 55.4 | 0.2 - 1058.9 | 57.2 ± 97.2 | 24.9 ± 54.6 | 0.2 - 1058.9 | 48.8 ± 60.3 | 24.8 ± 63.4 | 0.2 - 450.2 | 0.606 |
| 3MHA+4MHA | 425.3 ± 669 | 174.9 ± 501.2 | 3.4 - 9253.9 | **420.4 ± 687.6** | 158.3 ± 483.3 | 3.4 - 9253.9 | **478.4 ± 651.5** | 322.6 ± 548.7 | 3.4 - 4879.9 | **0.025** |

All the VOC metabolites were normalized by the urinary creatinine. Units: $\mu\text{g/g}$ creatinine.
SD: standard deviation, IQR: the interquartile range, min: minimum, max: maximum
Statistical tests for the comparison between nonsmokers and smokers: the t-test of log-transformed VOC metabolites. Bold values represent significant differences between these two subgroups.

**Supplemental Table S2A:**
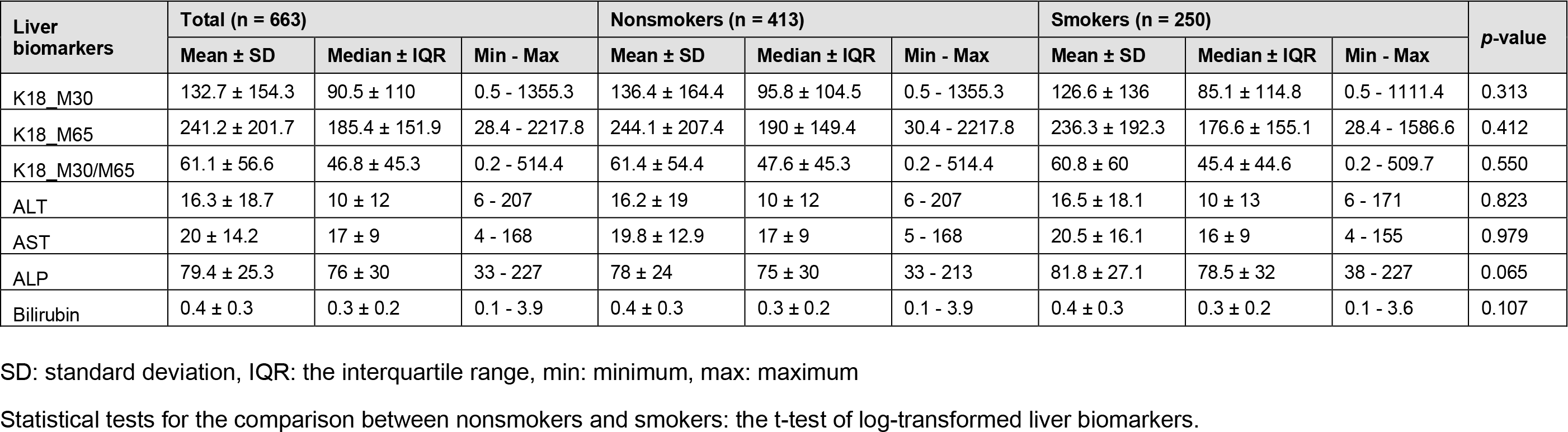
Distribution of serum keratin 18 (K18), liver enzymes and direct bilirubin within nonsmokers and smokers.

**Supplemental Table S2B:** Distribution of serum keratin 18 (K18), liver enzymes and direct bilirubin within males and females.

| Liver biomarkers | Total (n = 663) |  |  | Males (n = 262) |  |  | Females (n = 401) |  |  | p-value |
| --- | --- | --- | --- | --- | --- | --- | --- | --- | --- | --- |
| | Mean ± SD | Median ± IQR | Min - Max | Mean ± SD | Median ± IQR | Min - Max | Mean ± SD | Median ± IQR | Min - Max | |
| K18_M30 | 132.7 ± 154.3 | 90.5 ± 110 | 0.5 - 1355.3 | 135.9 ± 160 | 90.4 ± 109.2 | 0.5 - 1355.3 | 130.7 ± 150.5 | 90.7 ± 110.7 | 0.5 - 1355.3 | 0.230 |
| K18_M65 | 241.2 ± 201.7 | 185.4 ± 151.9 | 28.4 - 2217.8 | 251.7 ± 205.7 | 194.1 ± 161.7 | 30.4 - 2217.8 | 234.3 ± 199 | 181 ± 148.6 | 28.4 - 1870.5 | 0.077 |
| K18_M30/M65 | 61.1 ± 56.6 | 46.8 ± 45.3 | 0.2 - 514.4 | 58 ± 45.2 | 44.7 ± 43.8 | 0.3 - 322.1 | 63.2 ± 62.9 | 47.7 ± 45.6 | 0.2 - 514.4 | 0.885 |
| ALT | 16.3 ± 18.7 | 10 ± 12 | 6 - 207 | **21.2 ± 24.8** | 14 ± 18 | 6 - 207 | **13.1 ± 12.2** | 9 ± 9 | 6 - 124 | **<.0001** |
| AST | 20 ± 14.2 | 17 ± 9 | 4 - 168 | **23.4 ± 17.3** | 19 ± 9 | 6 - 168 | **17.8 ± 11.1** | 15 ± 8 | 4 - 128 | **<.0001** |
| ALP | 79.4 ± 25.3 | 76 ± 30 | 33 - 227 | 76.9 ± 23.2 | 74 ± 28 | 35 - 213 | 81.1 ± 26.5 | 77 ± 33 | 33 - 227 | 0.062 |
| Bilirubin | 0.4 ± 0.3 | 0.3 ± 0.2 | 0.1 - 3.9 | **0.5 ± 0.3** | 0.4 ± 0.2 | 0.1 - 3.9 | **0.4 ± 0.3** | 0.3 ± 0.2 | 0.1 - 3.6 | **<.0001** |

SD: standard deviation, IQR: the interquartile range, min: minimum, max: maximum
Statistical tests for the comparison between nonsmokers and smokers: the *t*-test of log-transformed liver biomarkers.

**Supplemental Table S2C:** Distribution of serum keratin 18 (K18), liver enzymes and direct bilirubin within White and Black persons.

| Liver biomarkers | Total (n = 663) |  |  | White (n = 514 ) |  |  | Black (n = 114) |  |  | p-value |
| --- | --- | --- | --- | --- | --- | --- | --- | --- | --- | --- |
| | Mean ± SD | Median ± IQR | Min - Max | Mean ± SD | Median ± IQR | Min - Max | Mean ± SD | Median ± IQR | Min - Max | |
| K18_M30 | 132.7 ± 154.3 | 90.5 ± 110 | 0.5 - 1355.3 | **141.6 ± 160** | 97.5 ± 116 | 0.5 - 1355.3 | **100.3 ± 139.7** | 60.8 ± 74.6 | 0.5 - 1111.4 | **<.0001** |
| K18_M65 | 241.2 ± 201.7 | 185.4 ± 151.9 | 28.4 - 2217.8 | **254.5 ± 208** | 194.6 ± 166 | 28.4 - 2217.8 | **197.8 ± 190.5** | 143.2 ± 128 | 30 - 1328.8 | **<.0001** |
| K18_M30/M65 | 61.1 ± 56.6 | 46.8 ± 45.3 | 0.2 - 514.4 | **63.1 ± 57** | 47.9 ± 44.9 | 0.2 - 514.4 | **52.4 ± 56.6** | 39.8 ± 36.1 | 0.3 - 509.7 | **0.012** |
| ALT | 16.3 ± 18.7 | 10 ± 12 | 6 - 207 | **17.1 ± 19.1** | 11 ± 14 | 6 - 207 | **14.1 ± 18.6** | 8.5 ± 7 | 6 - 171 | **0.014** |
| AST | 20 ± 14.2 | 17 ± 9 | 4 - 168 | 20.5 ± 14.1 | 17 ± 9 | 4 - 168 | 19.1 ± 16.1 | 15.5 ± 7 | 7 - 155 | 0.078 |
| ALP | 79.4 ± 25.3 | 76 ± 30 | 33 - 227 | 79.3 ± 24.7 | 76 ± 30 | 33 - 227 | 80.2 ± 27.7 | 74.5 ± 30 | 39 - 172 | 0.889 |
| Bilirubin | 0.4 ± 0.3 | 0.3 ± 0.2 | 0.1 - 3.9 | **0.4 ± 0.3** | 0.3 ± 0.3 | 0.1 - 3.9 | **0.3 ± 0.2** | 0.3 ± 0.2 | 0.1 - 1.2 | **<.0001** |

SD: standard deviation, IQR: the interquartile range, min: minimum, max: maximum
Statistical tests for the comparison between nonsmokers and smokers: the t-test of log-transformed liver biomarkers.

**Supplemental Table S3:** Assessment of smoking as a confounder for ALP associations in the total population (using cotinine as a categorical variable in our model). Six other confounders adjusted for include: age, sex, race, BMI, diabetes status, alcohol consumption.

|  |  | 6 adjustments |  |  |  | 7 adjustments |  |  |  |
| --- | --- | --- | --- | --- | --- | --- | --- | --- | --- |
| Enzymes_bilirubin | VOC metabolites | Percent_Change | UpperCL | LowerCL | p-value | Percent_Change | UpperCL | LowerCL | p-value |
| ALP | CEMA | **2.28** | **3.87** | **0.72** | **0.004** | 1.56 | 3.40 | -0.25 | 0.092 |
| ALP | 3HPMA | **1.78** | **3.17** | **0.40** | **0.011** | 0.88 | 2.72 | -0.93 | 0.343 |
| ALP | AAMA | **3.31** | **5.16** | **1.49** | **0.000** | **2.69** | **4.78** | **0.64** | **0.010** |
| ALP | CYMA | **0.86** | **1.53** | **0.18** | **0.012** | 0.25 | 1.40 | -0.89 | 0.666 |
| ALP | MU | 0.40 | 1.85 | -1.03 | 0.582 | -0.23 | 1.27 | -1.71 | 0.762 |
| ALP | DHBMA | 2.60 | 5.63 | -0.34 | 0.083 | 1.34 | 4.51 | -1.74 | 0.398 |
| ALP | MHBMA3 | **1.89** | **3.19** | **0.60** | **0.004** | 1.19 | 3.09 | -0.66 | 0.209 |
| ALP | HPMMA | **1.62** | **3.08** | **0.18** | **0.028** | 0.40 | 2.41 | -1.57 | 0.694 |
| ALP | AMCC | 1.21 | 2.88 | -0.43 | 0.150 | -0.54 | 1.63 | -2.66 | 0.622 |
| ALP | PGA | 1.72 | 3.74 | -0.27 | 0.091 | 0.71 | 2.88 | -1.42 | 0.517 |
| ALP | 2HPMA | 1.37 | 2.96 | -0.20 | 0.087 | 0.66 | 2.35 | -1.02 | 0.444 |
| ALP | PHEMA | 1.50 | 3.33 | -0.31 | 0.104 | 0.65 | 2.60 | -1.27 | 0.511 |
| ALP | MA | **3.40** | **5.83** | **1.03** | **0.005** | 2.21 | 5.07 | -0.57 | 0.120 |
| ALP | BMA | -1.08 | 0.79 | -2.91 | 0.255 | -1.10 | 0.75 | -2.92 | 0.243 |
| ALP | 2MHA | 0.50 | 1.45 | -0.43 | 0.293 | -0.19 | 0.88 | -1.25 | 0.729 |
| ALP | 3MHA+4MHA | **1.39** | **2.53** | **0.25** | **0.016** | 0.69 | 2.09 | -0.69 | 0.330 |

**Supplemental Table S4:** Assessment of smoking as a confounder for bilirubin associations in the total population (using cotinine as a categorical variable in our model. Six other confounders adjusted for include: age, sex, race, BMI, diabetes status, alcohol consumption).

| Enzymes_bilirubin | VOC metabolites | 6 adjustments |  |  |  | 7 adjustments |  |  |  |
| --- | --- | --- | --- | --- | --- | --- | --- | --- | --- |
|  |  | Percent_Change | UpperCL | LowerCL | p-value | Percent_Change | UpperCL | LowerCL | p-value |
| Bilirubin | CEMA | -0.83 | 1.80 | -3.40 | 0.532 | 0.36 | 3.49 | -2.67 | 0.818 |
| Bilirubin | 3HPMA | 0.90 | 3.26 | -1.42 | 0.451 | **3.84** | **7.07** | **0.70** | **0.016** |
| Bilirubin | AAMA | **-4.03** | **-1.08** | **-6.90** | **0.008** | **-3.74** | **-0.37** | **-6.99** | **0.030** |
| Bilirubin | CYMA | -1.01 | 0.12 | -2.13 | 0.080 | -0.79 | 1.15 | -2.70 | 0.422 |
| Bilirubin | MU | **3.75** | **6.28** | **1.27** | **0.003** | **4.82** | **7.49** | **2.21** | **0.000** |
| Bilirubin | DHBMA | **-5.97** | **-1.22** | **-10.48** | **0.014** | **-5.28** | **-0.19** | **-10.11** | **0.042** |
| Bilirubin | MHBMA3 | -0.46 | 1.72 | -2.60 | 0.676 | 1.73 | 4.99 | -1.43 | 0.288 |
| Bilirubin | HPMMA | -0.68 | 1.77 | -3.07 | 0.585 | 1.32 | 4.80 | -2.04 | 0.446 |
| Bilirubin | AMCC | **-3.73** | **-1.03** | **-6.35** | **0.007** | **-4.01** | **-0.43** | **-7.46** | **0.028** |
| Bilirubin | PGA | -3.05 | 0.25 | -6.25 | 0.070 | -2.36 | 1.27 | -5.85 | 0.200 |
| Bilirubin | 2HPMA | -0.73 | 1.93 | -3.33 | 0.586 | 0.10 | 3.00 | -2.71 | 0.943 |
| Bilirubin | PHEMA | -2.80 | 0.20 | -5.71 | 0.067 | -2.22 | 1.03 | -5.36 | 0.178 |
| Bilirubin | MA | -1.87 | 2.10 | -5.69 | 0.351 | -0.16 | 4.66 | -4.77 | 0.945 |
| Bilirubin | BMA | 0.27 | 3.50 | -2.87 | 0.869 | 0.29 | 3.52 | -2.84 | 0.857 |
| Bilirubin | 2MHA | -0.72 | 0.88 | -2.28 | 0.376 | -0.11 | 1.72 | -1.91 | 0.906 |
| Bilirubin | 3MHA+4MHA | **-1.96** | **-0.07** | **-3.81** | **0.043** | -1.54 | 0.80 | -3.82 | 0.196 |

**Supplemental Figure S1:**
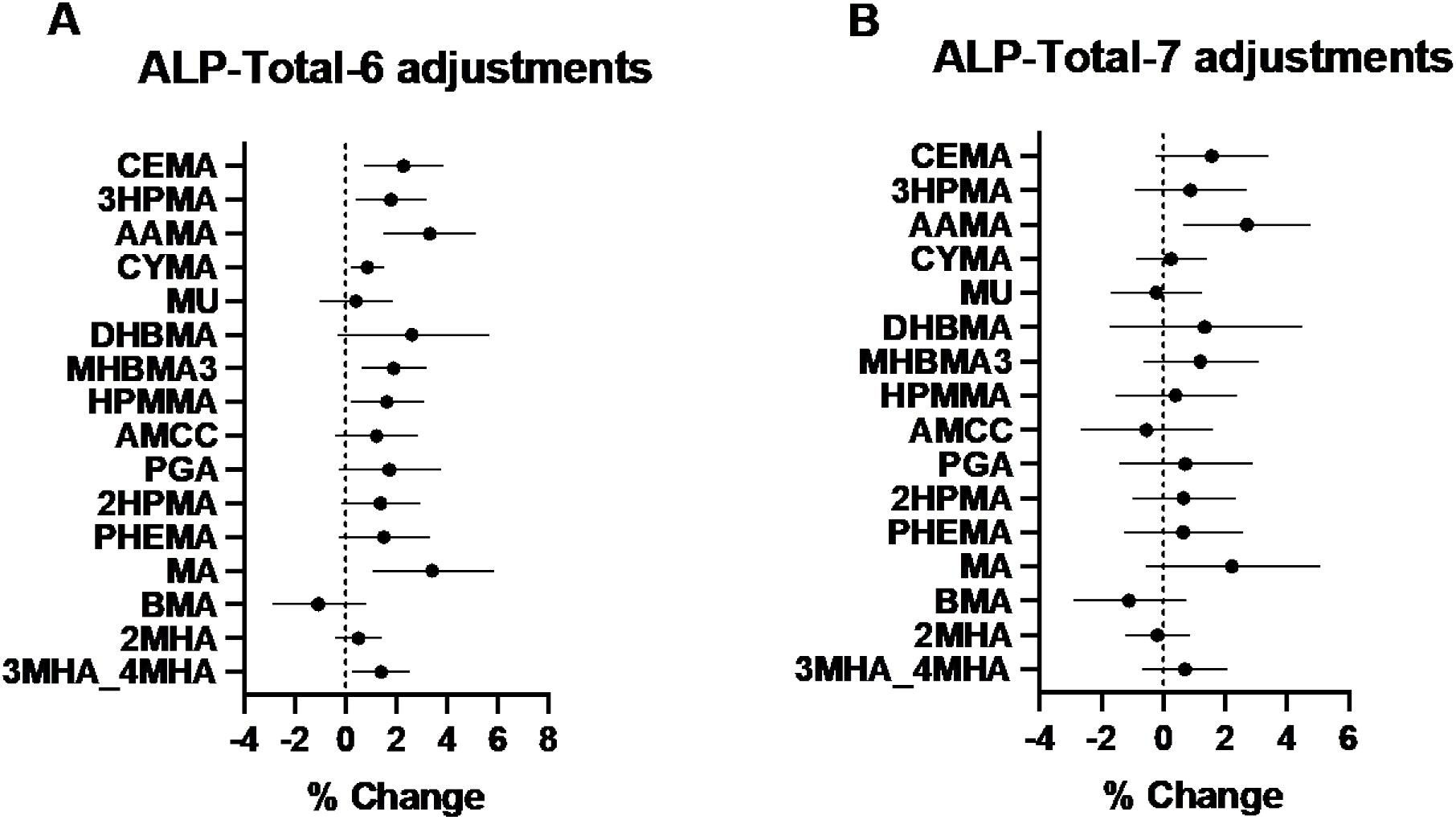
Comparison of adjusted associations between urinary levels of 16 VOC metabolites and ALP in the total study population (n=663) between 6 adjustments and 7 adjustments (6 adjustments + smoking). Represents % change (95% CI) per 2-fold increase in urinary VOC metabolite levels. Generalized linear models were used to examine the adjusted associations. A. Models were adjusted for 6 common liver-related confounders: age, sex, race, BMI, diabetes status, and alcohol consumption. B. Models were adjusted for 6 common liver-related confounders and smoking. The statistical significance was set at the p-value <0.05.

**Supplemental Figure S2:**
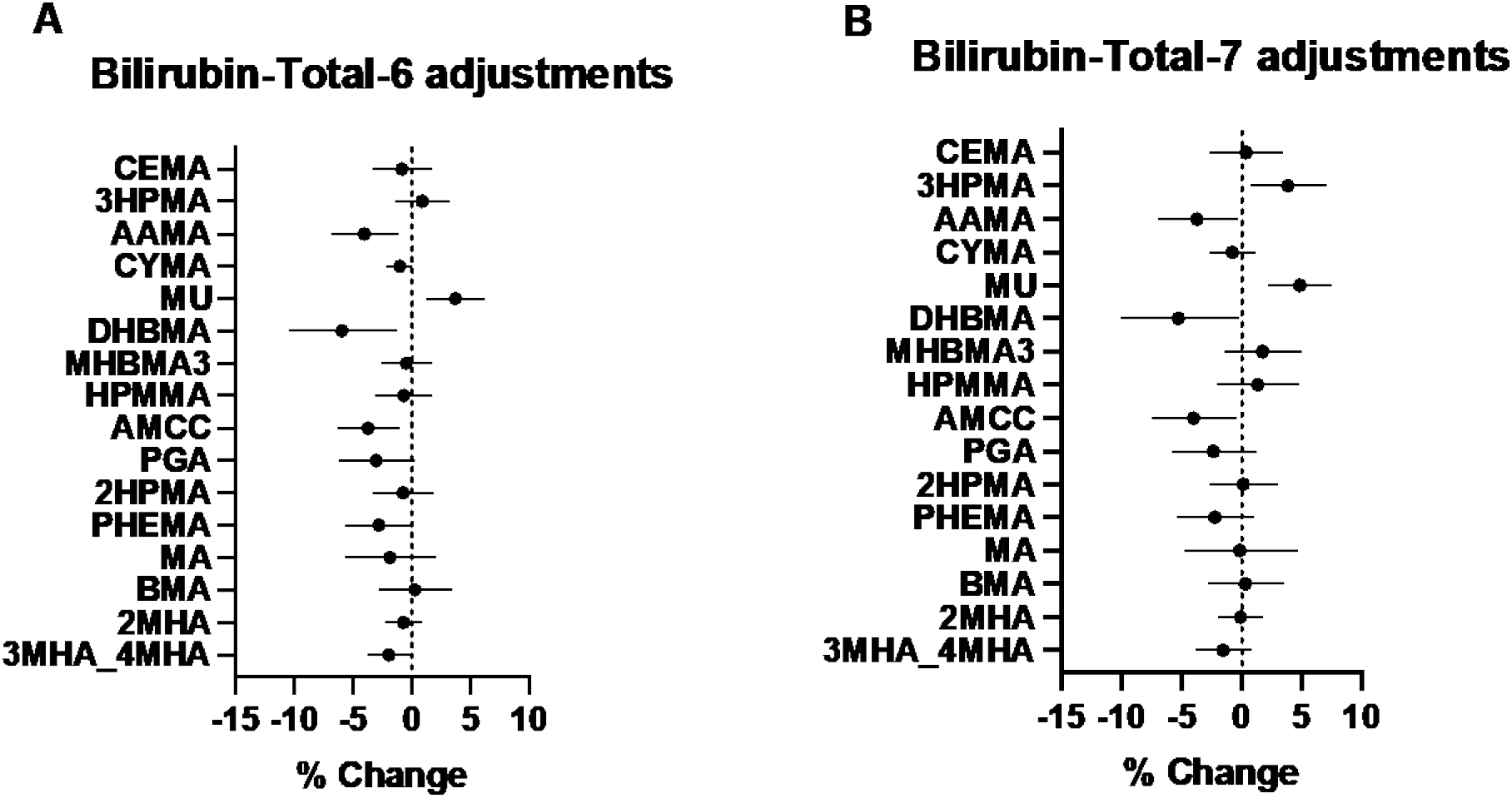
Comparison of adjusted associations between urinary levels of 16 VOC metabolites and Bilirubin in the total study population (n=663) between 6 adjustments and 7 adjustments (6 adjustments + smoking). Represents % change (95% CI) per 2-fold increase in urinary VOC metabolite levels. Generalized linear models were used to examine the adjusted associations. A. Models were adjusted for 6 common liver-related confounders: age, sex, race, BMI, diabetes status, and alcohol consumption. B. Models were adjusted for 6 common liver-related confounders and smoking. The statistical significance was set at the p-value <0.05.

## Notes

### Competing Interest Statement

The authors have declared no competing interest.

### Author Declarations

The study was approved by the University of Louisville Institutional Review Board (IRB 15.1260).

